# The ADNEX risk prediction model for ovarian cancer diagnosis: A systematic review and meta-analysis of external validation studies

**DOI:** 10.1101/2023.07.12.23291935

**Authors:** Lasai BARREÑADA, Ashleigh LEDGER, Paula DHIMAN, Gary S. COLLINS, Laure WYNANTS, Jan Y. VERBAKEL, Dirk TIMMERMAN, Lil VALENTIN, Ben VAN CALSTER

## Abstract

Objectives: To conduct a systematic review of studies externally validating the ADNEX model for ovarian cancer diagnosis and perform a meta-analysis of its performance. Design: Systematic review, meta-analysis Data sources: Medline, EMBASE, WOS, Scopus, and EuropePMC up to 15/05/2023. Review methods: We included external validation studies of the performance of ADNEX using any study design and any study population comprising patients with an adnexal mass. Two independent reviewers extracted data. Disagreements were resolved through discussion. Reporting quality of the studies was scored using the TRIPOD reporting guideline and methodological conduct and risk of bias using the PROBAST tool. We performed random effects meta-analysis of the AUC, sensitivity and specificity at the 10% risk of malignancy threshold, and Net Benefit and Relative Utility at the 10% risk of malignancy threshold. Results: We included 47 studies (17,007 tumours) with median study sample size 261 (range 24-4905). On average, 61% of TRIPOD items were reported. Handling of missing data, sample size justification, and model calibration were rarely described. 91% of validations were at high risk of bias, mainly due to the unexplained exclusion of incomplete cases, low sample size, or absent calibration assessment. The summary AUC to distinguish benign from malignant tumours in operated patients was 0.93 (95% CI 0.92-0.94, 95% prediction interval 0.85-0.98) for ADNEX with CA125 as a predictor (9202 tumours, 43 centres, 18 countries, 21 studies) and 0.93 (95% CI 0.91-0.94, 95% prediction interval 0.85-0.98) for ADNEX without CA125 (6309 tumours, 31 centres, 13 countries, 12 studies). The estimated probability that the model has clinical utility in a new centre was 95% (with CA125) and 91% (without CA125). When restricting analysis to studies at low risk of bias, summary AUCs were 0.93 (with CA125) and 0.91 (without CA125), and estimated probabilities that the model has clinical utility were 89% (with CA125) and 87% (without CA125). Discussion: ADNEX performed well to distinguish benign from malignant tumours in populations from different countries and settings regardless of whether CA125 was used or not. A key limitation is that calibration was rarely assessed. Review registration: PROSPERO, CRD42022373182

## INTRODUCTION

The optimal management of patients with an ovarian mass depends on the histology of the mass. Patients with a benign mass can be managed non-surgically with clinical and ultrasound follow-up or using conservative surgical techniques.[1,2] Malignant tumours benefit from management in specialised oncology centres, but borderline malignancies, stage I primary invasive tumours, and advanced primary invasive tumours may require different surgical approaches.[3,4] To optimise patient triage without operating on all masses, diagnostic models can be used to estimate the likelihood of malignancy and so be used to plan treatment for patients.

Given the potential advantages of accurately predicting risk of malignancy, the International Ovarian Tumour Analysis (IOTA) group developed the Assessment of Different NEoplasias in the adneXa (ADNEX) risk prediction model, using three clinical and six ultrasound predictor variables.[5] The clinical variables are age, serum CA-125 level, and type of centre (oncology centre vs other). An oncology centre is defined as a tertiary referral centre with a specific gynaecology oncology unit. The ultrasound variables are the maximal diameter of the lesion, proportion of solid tissue (defined as the largest diameter of the largest solid component divided by the largest diameter of the lesion), number of papillary projections, presence of more than 10 cyst locules, presence of acoustic shadows, and ascites. ADNEX is a multinomial logistic regression model that estimates the risk of five tumour types: benign, borderline, stage I primary invasive, stage II-IV primary invasive, and secondary metastatic. The total risk of malignancy calculated by ADNEX is the sum of the risks for each malignant subtype. ADNEX has two versions: one with and one without CA125 as a predictor.[5,6] The model was developed on data from 5909 patients with an adnexal mass that subsequently underwent surgery and that were recruited at 24 centres across 10 countries (Belgium, Italy, Czech Republic, Poland, Sweden, China, France, Spain, UK and Canada). Although developed on data from patients that underwent surgery, ADNEX has been validated on cohorts that also include non-surgically managed patients.[7–10]

ADNEX is widely used and is included in national guidelines, e.g. in Belgium, The Netherlands and Sweden.[11–13] It is recommended by scientific societies, such as International Society of Ultrasound in Obstetrics and Gynecology (ISUOG), European Society of Gynaecological Oncology (ESGO), European Society for Gynaecological Endoscopy (ESGE), and the American College of Radiology.[4,14] In addition, manufacturers of ultrasound machines have begun to incorporate ADNEX directly into their machines.[15–17]

Several external validation studies of ADNEX have been carried out.[8–10,18–61] To date, five published systematic reviews and meta-analyses of ADNEX have summarised between 3 and 22 external validation studies.[62–67] All systematic reviews evaluated ADNEX solely as a diagnostic test, reporting a summary sensitivity and specificity at a threshold for the estimated risk of malignancy of 10% or 15%.[62,64,65,67,68] However, the ADNEX model is not only a diagnostic test. It is also a risk prediction model to provide probability estimates of five different tumour types at the individual patient level. Using classification thresholds reduces the information given by the model.[69] Further, when analysing ADNEX merely as a diagnostic test with a 10% threshold we assume that there is no difference between patients with an estimated risk of 11% versus 99% of having a malignancy. This means that these meta-analyses have not fully validated the diagnostic performance of ADNEX. For example, pooling discrimination performance (area under the receiver operating characteristic curve, AUC) allows us to determine the ability of the model to differentiate between patients with and without the outcome across thresholds. There are now guidelines on how to evaluate the quality and risk of bias of external validation studies of risk prediction models. [70–72] These should be used in meta-analyses of validation studies.

The objective of this study is to conduct a systematic review of studies that externally validate ADNEX, to describe reporting completeness and risk of bias of the validation studies, and to conduct meta-analyses of model performance measures.

## METHODS

### Protocol registration

We report this study according to the PRISMA (Preferred Reporting Items for Systematic reviews and Meta-Analysis) and TRIPOD-SRMA (Transparent Reporting of multivariable prediction models for Individual Prognosis Or Diagnosis: checklist for Systematic Reviews and Meta-Analyses) checklists.[72,73] The study protocol was prospectively registered in the International prospective register of systematic reviews (PROSPERO; ID CRD42022373182).[74]

### Eligibility criteria

Any study that carried out an external validation to evaluate the performance of the ADNEX model using any study design and any study population was eligible for inclusion in the systematic review.

The exclusion criteria were (1) studies that did not evaluate the model performance of ADNEX in any way; (2) studies that evaluated the predictive performance only of updated versions of ADNEX; (3) studies for which only an abstract is available, or the full text cannot be obtained; (4) case studies presenting ADNEX performance for individual patients (this criterion was not pre-specified in the protocol but was added post hoc upon review of the search results). Updating can refer to recalibration, refitting, or extension with additional predictors.[75]

### Information sources and search strategy

We created a search string and overall search strategy with the help of biomedical reference librarians of the KU Leuven Libraries. We searched the electronic databases Medline (via PubMed), EMBASE, Web of Science, and Scopus for published articles, and EuropePMC for preprints. The search dates were from the publication of the first ADNEX paper (15/10/2014) until 15/05/2023 (date the final search was run). We also screened all articles citing the original ADNEX paper.[5] The reference lists of relevant review and opinion articles retrieved by the search strings were checked for other potentially eligible articles. Forward and backward snowballing (forward/back cross-reference checking) of the included articles was performed to identify additional publications.[76] There was no language restriction, but for papers in languages other than English, Spanish, Dutch, French or Swedish an automatic translation tool (deepl.com) was used to decide whether to include or exclude a paper and to extract information. The full search strategy is provided in **Supplementary Material S1**.

### Study selection

The studies we identified in our search were imported into Zotero reference manager, where they were automatically de-duplicated. The de-duplicated records were then imported into Rayyan web application for manual de-duplication (LB) and subsequent screening of title and abstract by two independent authors with any conflicts being resolved by discussions between them (LB, AL).[77]

As three authors (BVC, LV, DT) are members of the IOTA group that developed ADNEX, we separated the studies as linked or not linked to IOTA. A study was IOTA-linked if it was co-authored by a member of the IOTA steering committee (**Supplementary Material S2**). IOTA-linked papers, as well as a few others with a potential conflict of interest (i.e., including authors that are or were IOTA collaborators), were independently assessed by authors PD and GSC, two medical statisticians with expertise in prediction modelling and unrelated to IOTA. All other studies were independently assessed by authors LB and AL. Conflicts were resolved through discussion between reviewers and for the non-IOTA papers also through discussion with authors BVC, LV and JYV.

### Data extraction and data items

Data was extracted and entered into a standardised data extraction form in Microsoft Excel. The data extraction focused on general and design characteristics of studies, target population, reference standard, sample size, performance results, reporting quality, methodological quality, and risk of bias (**Table S1**). The extraction form was based on and adapted from the CHARMS (CHecklist for critical Appraisal and data extraction for systematic Reviews of prediction Modelling Studies), TRIPOD (Transparent Reporting of a multivariable prediction model for Individual Prognosis Or Diagnosis), and PROBAST (Prediction model Risk Of Bias ASsessment Tool) tools.[71,78,79]

To describe model performance, we extracted information on any reported measure related to discrimination, calibration, diagnostic accuracy, or clinical utility. The reference standard could be binary (e.g., benign vs malignant) or multinomial (e.g., the five tumour types predicted by ADNEX). Performance data was extracted for all reported validations (i.e., for both ADNEX versions), subgroup analyses, sensitivity analyses, and centre-specific results in multicentre studies.

For each study we assessed the reporting of all TRIPOD items that are applicable to external validation studies (**Table S2**). We also checked PROBAST’s ‘signalling questions’ and evaluated risk of bias for each subdomain (participants, predictors, outcome, and analysis) and overall. We included rationale for the risk of bias classification.

We contacted study authors to obtain further information or results in the following situations: (1) when centre-specific results were not reported in multicentre studies, (2) the type of centre was not explicitly reported (in case of nonresponse, the clinical co-authors, JYV, DT, and LV, classified the centre), (3) overall performance was reported but not performance by menopausal status, or (4) performance was not reported for operated patients separately in studies including both surgically and non-surgically managed patients.

Details on all extracted items are available in **Supplementary Material (Table S1)** and **Open Science Framework repository (Extraction sheet)**.[80]

### Statistical analysis and quantitative data synthesis

Data were summarised with descriptive statistics and data visualisations. Meta-analysis of performance, using centre-specific results for multicentre studies where possible, was performed using random effects meta-analysis methods. Meta-analysis was done separately for the two versions of ADNEX. In addition to 95% confidence intervals (CI) for the summary performance, we assessed heterogeneity using tau squared and 95% prediction intervals (PI). **Supplementary Material S3-5** provides details on the meta-analysis methodology.

Meta-analysis of the AUC for benign versus malignant tumours was done on the logit scale. Meta-analysis for sensitivity and specificity was also performed on the logit scale using random effects meta-analysis.[81] As the meta-analysis was only conducted for the 10% threshold for the risk of malignancy, we did not use the bivariate random effects model as specified in the protocol.[74] Meta-analysis of Net Benefit (NB) [82,83] and Relative Utility (RU) [84,85] at the 10% risk of malignancy threshold was performed using Bayesian trivariate random effects meta-analysis of sensitivity, specificity and prevalence of malignancy.[86] For Bayesian methods, 95% credible intervals (CrI) are reported instead of 95% CIs. The Bayesian approach allowed to estimate the probability that the model is useful in a new centre, i.e. the probability that RU is >0.

To address multinomial discrimination performance, meta-analysis of AUCs between pairs of tumour outcomes (‘pairwise AUCs’) was conducted on logit scale. We only included studies that used the “conditional risk method” to calculate pairwise AUCs.[87]

Subgroups are defined based on geographical location, type of centre, and menopausal status. Sensitivity analysis are based on the risk of bias judgment and on whether the study was IOTA linked or not. As pre-specified in the protocol, we only meta-analysed performance if at least three estimates in a specific analysis could be retrieved from the included studies.[74] To assess the association of prevalence of malignancy with the AUC and sensitivity and specificity at the 10% risk of malignancy threshold, we used meta-regression.[88]

Reporting bias and small study effects were visually explored using funnel plots adapted for the AUC. The body of evidence was assessed using an adapted version of GRADE.[89]

All analyses were performed in R version 4.2.2 using package “metamisc” for the AUC, “meta” and “mada” for sensitivity and specificity, and rjags for NB and RU. [89–95] Bayesian methods were computed using JAGS version 4.3.1.[95]

## RESULTS

We identified 1843 records and screened 490 after de-duplication. Forty-seven studies met our inclusion criteria and were included in this systematic review **(Figure 1**, **Table S3**).[8–10,18–61] Three studies were excluded because they used the same data as in another included study, and one study was excluded because a preliminary version of ADNEX was used. [96–99] The data of three studies that were IOTA-linked and three other studies with a potential conflict of interest were extracted by authors PD and GSC.[7,27,40,49,51,52]

**Figure 1.**
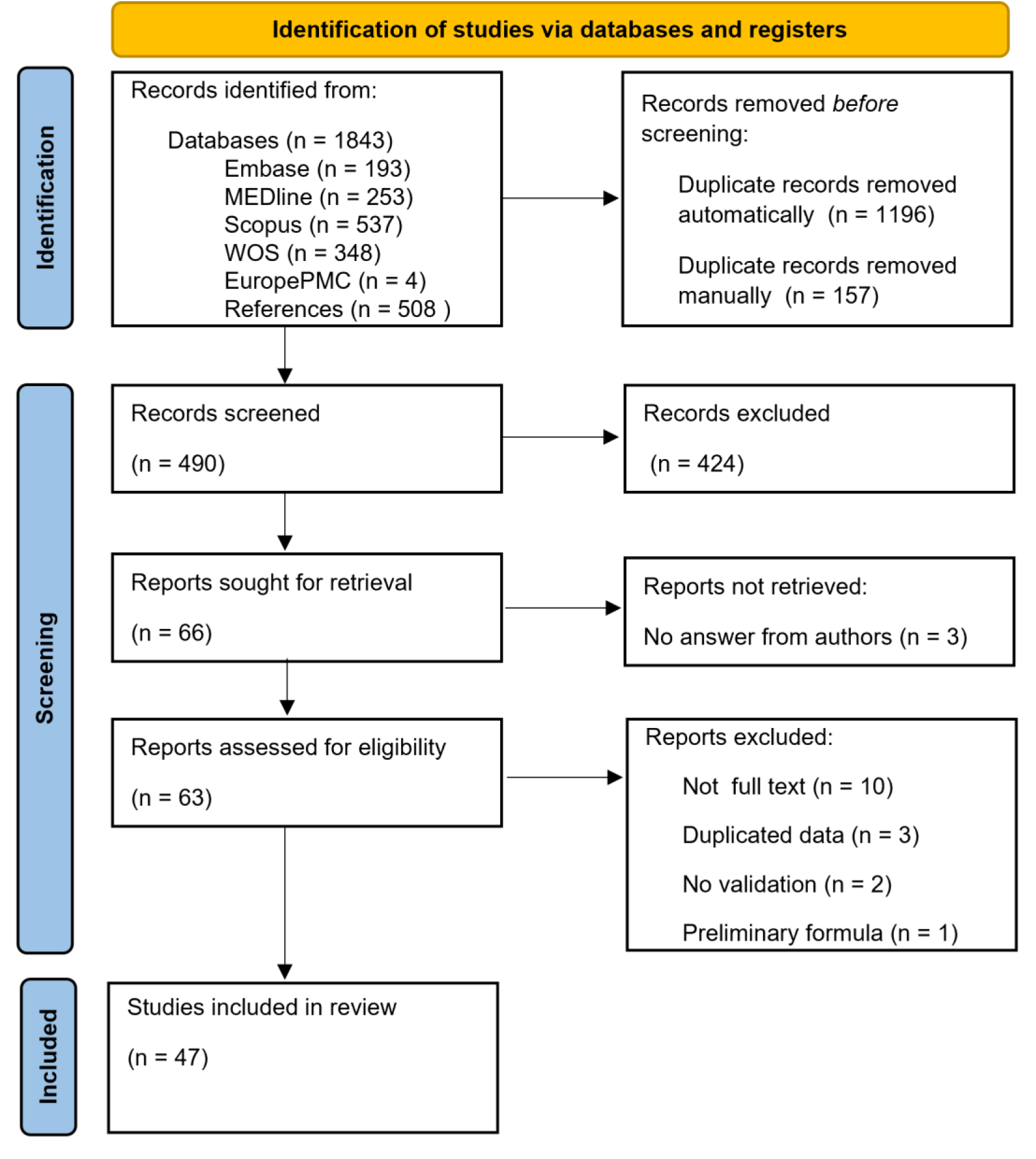
PRISMA (preferred reporting items for systematic reviews and meta-analyses) flowchart of study inclusions and exclusions.

Key study characteristics are summarised in **Table 1** (**Table S3** shows study-specific details). The unit of analysis was the patient in 42 (89%) studies and the tumour in five (11%) studies. When tumour was the unit of analysis, multiple tumours for the same patient could be included. The 47 studies reported on 17007 tumours, with a median study sample size of 261 tumours (range 24 to 4905). The validations were conducted in 28 countries with most studies being conducted in Asia (51%) and Europe (38%). **Supplementary Material S6** presents a list of reporting inconsistencies.

**Table 1.** Characteristics of the 47 included studies.

| **Study characteristic** | **N (%)** | **Comments** |
| --- | --- | --- |
| Unit of analysis |  |  |
| Patient | 42 (89) | 1 tumour per patient |
| Tumour | 5 (11) | >1 tumour per patient possible |
| Region |  |  |
| Asia | 24 (51) | Common countries: China (12), South Korea (3) |
| Europe | 18 (38) | Common countries: Poland (7), Italy (4), Spain (4), UK (3), Sweden (3) |
| South America | 3 (6) |  |
| North America | 2 (4) |  |
| Number of centres |  |  |
| 1 | 37 (79) |  |
| 2-5 | 7 (15) |  |
| >5 | 3 (6) | Range: 8-17 |
| Type of centre |  |  |
| Oncology centre(s) | 35 (74) |  |
| Non-oncology centre(s) | 2 (4) |  |
| Both types of centres | 4 (9) |  |
| Unclear | 6 (13) |  |
| ADNEX version |  |  |
| ADNEX with CA125 | 37 (79) | 21 only used ADNEX with CA125, 16 used both |
| ADNEX without CA125 | 19 (40) | 3 only used ADNEX without CA125, 16 used both |
| Mixed | 3 (6) | ADNEX with CA125 used if CA125 was available |
| Unclear | 4 (9) |  |
| Selection based on histology |  |  |
| No | 39 (83) |  |
| Yes | 8 (17) | e.g., borderlines excluded, only invasive tumours |
| Focus only on clinical subgroup |  |  |
| No | 42 (89) |  |
| Yes | 5 (11) | e.g., only pregnant patients |
| Target population |  |  |
| Surgically managed patients | 43 (91) |  |
| Surgically and non-surgically managed patients | 4 (9) |  |

ADNEX with CA125 was validated in 37 (79%) studies, and ADNEX without CA125 in 19 (40%) studies (16 studies evaluated both versions). Three (6%) studies conducted a mixed validation, using ADNEX with CA125 when CA125 was available. For four (9%) studies, the ADNEX version was unclear. In total, 63 validations of ADNEX were performed after distinguishing between the ADNEX versions used (**Table S4**). When also including reported results for subgroups (e.g., by menopausal status, or by centre in multicentre studies), the total number of validations reported across the 47 studies was 159.

Five (11%) studies focused only on a specific clinical subgroup, such as pregnant women or tumours for which the clinician’s subjective assessment of the outcome was uncertain.[10,24,38,42,52] Eight (17%) studies selected patients based on histology (**Table S3** for details). Thirty-six (77%) studies did not focus on a specific clinical subgroup and did not select tumours based on histology. In these 36 studies that were eligible for the meta-analysis, the median sample size was 284 tumours (range 50-4905), the median number of malignant tumours was 68 (7-1041) and the median prevalence of malignancy 28% (3%-57%). Fourteen of the 36 (39%) studies had at least 100 benign and at least 100 malignant tumours.

The target population of the studies could be surgically managed patients or surgically and non surgically managed patients. The reference standard for determining the tumour type in surgically managed patients was always histopathology. Four (9%) studies included surgically and non-surgically managed patients. In non-surgically managed patients, the outcome determination was the clinician’s subjective assessment of the tumour as benign or malignant or spontaneous resolution of the tumour during follow-up. The required follow-up time to determine the outcome was 3-4 months, 1 year, or 2 years, depending on the study.[8–10,56]

The most commonly reported performance measure was the AUC for benign vs malignant tumours (72%) (**Table S5**). About two-thirds of studies (66%) presented a receiver operating characteristic curve, 31 (66%) reported sensitivity and specificity performance at the 10% threshold for the risk of malignancy, 12 (26%) reported measures for multinomial discrimination and four (9%) studies reported calibration performance.

### Critical appraisal: reporting and risk of bias

Completeness of reporting the TRIPOD items was assessed for the 63 validations. Adherence to TRIPOD items was on average 61%: studies reported on average 16.5 out of 27 items (**Figures 2 and S1**). The least commonly reported items were ‘comparison of demographics, predictors and outcome between the model development and external validation data’ (item 13c; 5%), ‘the reporting of performance measures with confidence intervals’ (item 16; 11%), ‘specification of all performance measures’ (item 10d; 11%), ‘rationale for study sample size’ (item 8; 13%), and ‘description of how missing data were handled’ (item 9; 22%).

**Figure 2.**
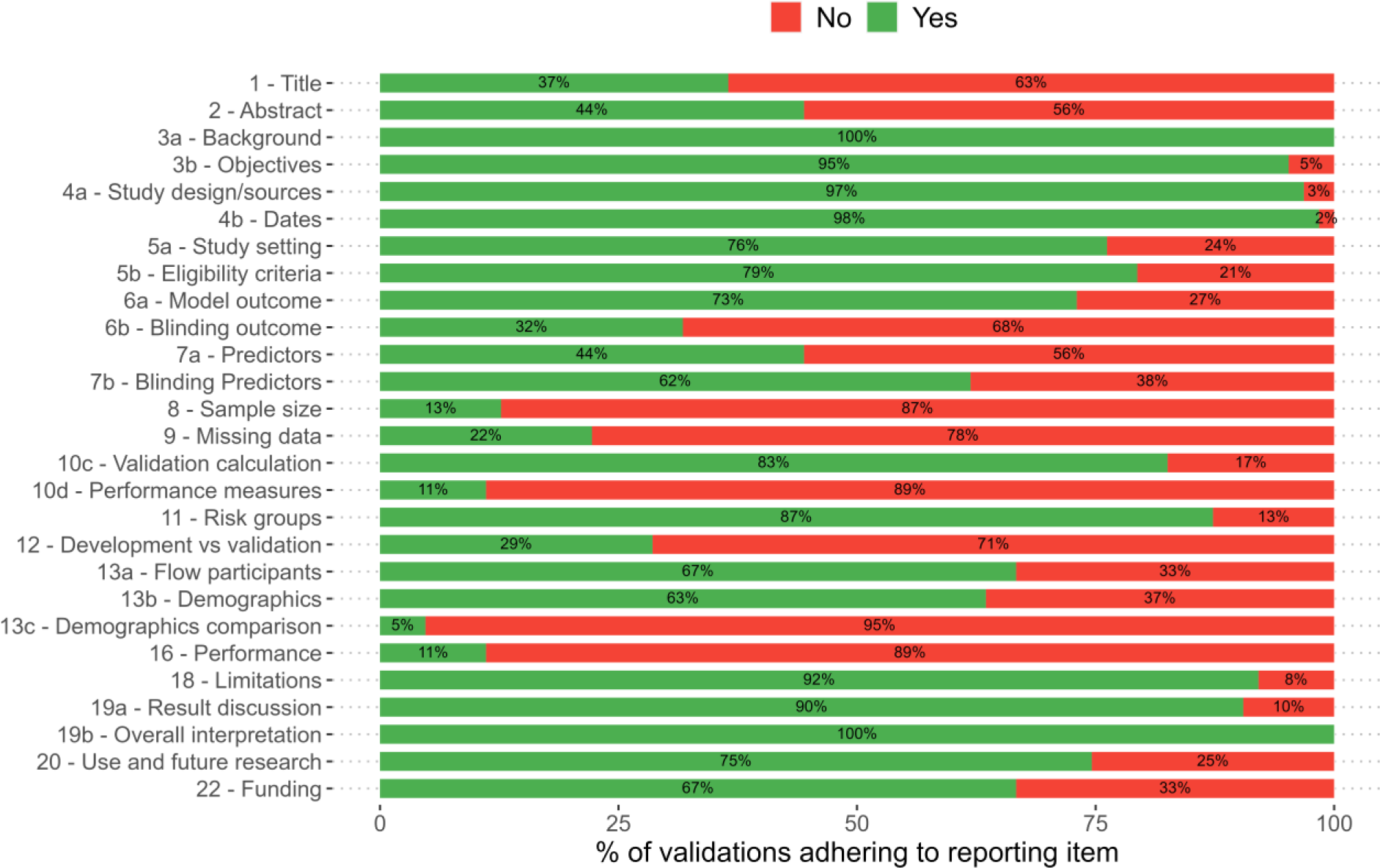
TRIPOD adherence per item in 63 validations.

Fifty-seven (90%) of 63 validations were rated at high risk of bias, two (3%) at uncertain risk of bias, and four (6%) at low risk of bias (**Figures 3, S2-5**,**OSF Extraction sheet**). 43 (68%) validations were at high risk of bias for the participant domain, mostly by having incomplete data as an exclusion criterion. Fifty-seven (90%) validations were at high risk of bias for the analysis domain, mostly due to small sample size (69%), not including all participants in the analysis (85%), inappropriate handling of missing data (82%), and incomplete evaluation of model performance - in most instances by not reporting an assessment of calibration (89%).

**Figure 3.**
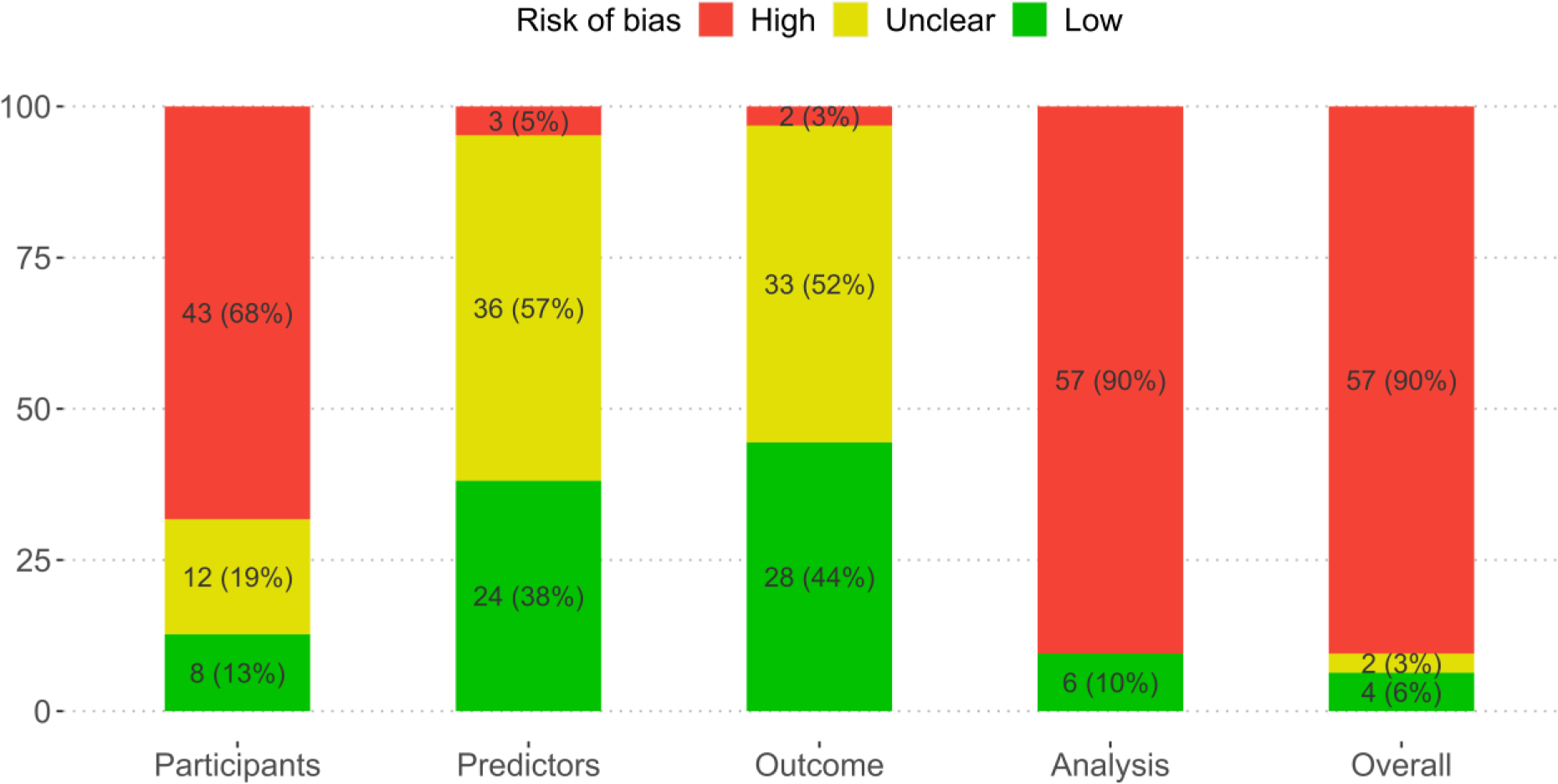
PROBAST risk of bias by subdomain and overall in 63 validations.

TRIPOD adherence in 36 studies without focus on selected histologies or clinical subgroups was on average 65% (17.47 items out of 27). On these studies, two where at low risk of bias, one at unclear risk of bias and 33 at high risk of bias.

### Meta-analysis

The meta-analysis included studies without post hoc selection based on histology and without focus on a clinical subgroup only (n = 36) that reported the meta-analysed metrics.

The AUC for benign vs malignant tumours in operated patients was reported in 12 studies (6309 tumours, 31 centres, 13 countries) for ADNEX without CA125 (**Table S4 and S6**), and the summary AUC was 0.93 (CI 95% 0.91-0.94, 95% PI 0.85-0.98) (**Table 2**, **Figure 4**).[22,26,30,31,37,43–46,49,56,61] Twenty-one studies (9202 tumours, 43 centres, 18 countries) reported the AUC for benign vs malignant tumours in operated patients for ADNEX with CA125,[18,19,23,26,30,35,37,40,41,44–46,49,51,52,55,56,58,60,61,100] (**Table S4 and S6**) and the summary AUC was 0.93 (CI 95% 0.92-0.94, 95% PI 0.85-0.98) (**Table 2**, **Figure 5**).

**Figure 4.**
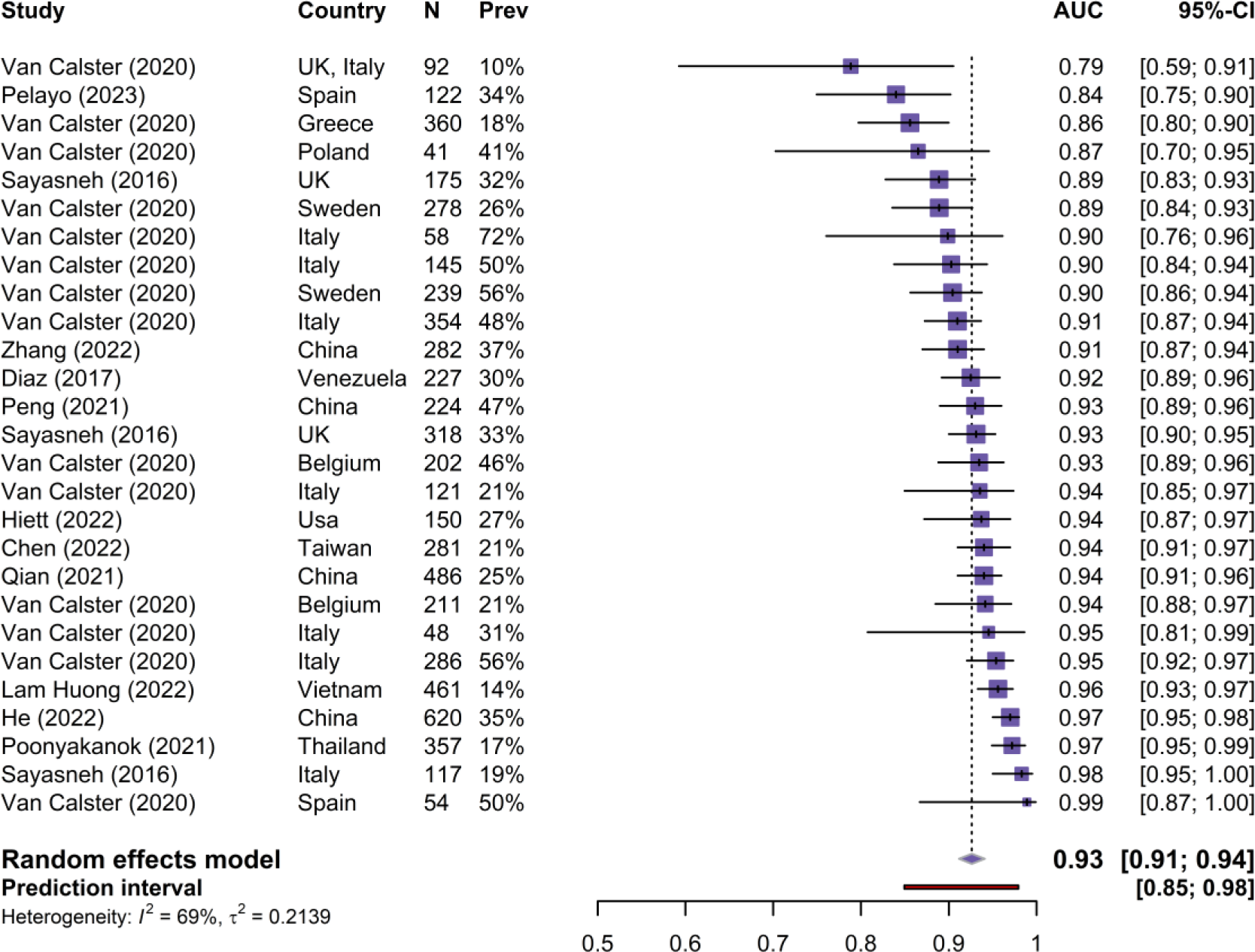
Forest plot of area under the receiver operating curve (AUC) for ADNEX without CA125. CI, confidence interval; Prev, prevalence of malignancy.

**Figure 5.**
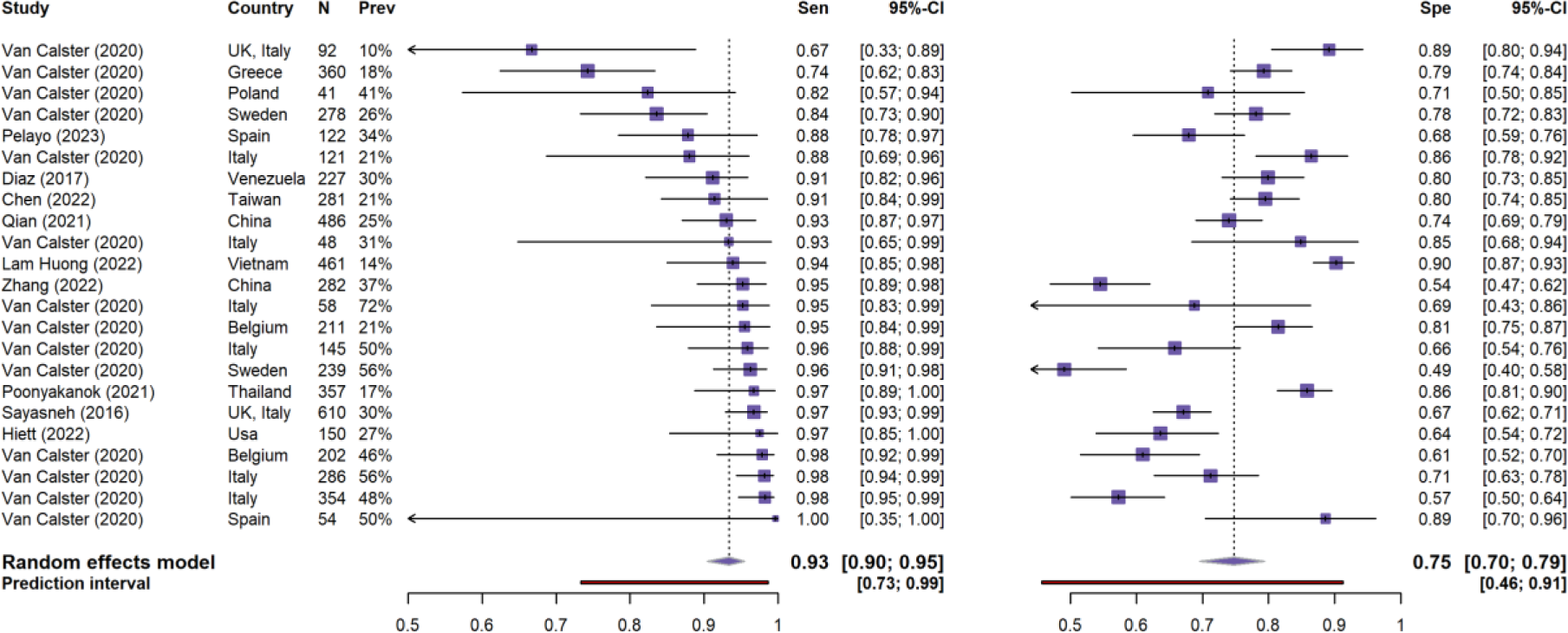
Forest plot of area under the receiver operating characteristic curve (AUC) for ADNEX with CA125. CI, confidence interval; Prev, prevalence of malignancy.

**Table 2.** Meta-analysis of the AUC to discriminate between benign and malignant tumours, including sensitivity and subgroup analyses.

| Meta-analysis | Number<br>of studies<br>(centres) | Summary estimate<br>(95% CI) | 95% PI | $\tau^2$ |
| --- | --- | --- | --- | --- |
| *Main analysis* |  |  |  |  |
| Operated patients, with CA125 | 21 (43) | 0.93 (0.92-0.94) | 0.85 to 0.98 | 0.25 |
| Operated patients, without CA125 | 12 (31) | 0.93 (0.91-0.94) | 0.85 to 0.98 | 0.21 |
| *Sensitivity analyses (operated patients)* |  |  |  |  |
| High or Unclear risk of bias studies, with CA125 | 19 (23) | 0.93 (0.91-0.95) | 0.84 to 0.99 | 0.33 |
| Low risk of bias studies, with CA125 | 2 (20) | 0.93 (0.91-0.94) | 0.87 to 0.97 | 0.12 |
| IOTA studies, with CA125 | 4 (23) | 0.92 (0.91-0.94) | 0.88 to 0.97 | 0.09 |
| Non-IOTA studies, with CA125 | 17 (20) | 0.93 (0.91-0.95) | 0.83 to 0.99 | 0.38 |
| High or Unclear risk of bias studies, without CA125 | 10 (11) | 0.94 (0.92-0.96) | 0.86 to 0.99 | 0.26 |
| Low risk of bias studies, without CA125 | 2 (20) | 0.91 (0.90-0.93) | 0.85 to 0.96 | 0.11 |
| IOTA studies, without CA125 | 2 (20) | 0.91 (0.89-0.93) | 0.85 to 0.97 | 0.14 |
| Non-IOTA studies, without CA125 | 10 (11) | 0.94 (0.92-0.96) | 0.86 to 0.99 | 0.26 |
| *Subgroup analyses (operated patients)* |  |  |  |  |
| Asian centres, with CA125 | 11 (13) | 0.94 (0.91-0.96) | 0.81 to 1 | 0.54 |
| Asian centres, without CA125 | 7 (8) | 0.95 (0.93-0.97) | 0.89 to 0.99 | 0.19 |
| Chinese centres, with CA125 | 5 (5) | 0.94 (0.91-0.97) | 0.87 to 0.99 | 0.25 |
| Chinese centres, without CA125 | 4 (4) | 0.94 (0.90-0.97) | 0.85 to 0.99 | 0.33 |
| European centres, with CA125 | 8 (28) | 0.92 (0.91-0.94) | 0.88 to 0.96 | 0.07 |
| European centres, without CA125 | 3 (21) | 0.91 (0.89-0.93) | 0.84 to 0.97 | 0.16 |
| Non-oncology centres, with CA125 | 2 (9) | 0.91 (0.88-0.94) | 0.85 to 0.97 | 0.11 |
| Non-oncology centres, without CA125 | 1 (8) | 0.90 (0.86-0.95) | 0.80 to 0.99 | 0.24 |
| Oncology centres, with CA125 | 18 (31) | 0.93 (0.92-0.95) | 0.84 to 0.98 | 0.29 |
| Oncology centres, without CA125 | 12 (23) | 0.93 (0.91-0.94) | 0.85 to 0.98 | 0.22 |
| Postmenopausal patients, with CA125 | 11 (32) | 0.92 (0.90-0.94) | 0.83 to 0.98 | 0.25 |
| Postmenopausal patients, without CA125 | 4 (22) | 0.90 (0.87-0.93) | 0.81 to 0.97 | 0.19 |
| Premenopausal patients, with CA125 | 11 (32) | 0.91 (0.89-0.94) | 0.81 to 0.98 | 0.28 |
| Premenopausal patients, without CA125 | 4 (22) | 0.91 (0.89-0.93) | 0.85 to 0.96 | 0.08 |
| *Target population* |  |  |  |  |
| Operated and non-surgically managed patients, with CA125 | 2 (18) | 0.94 (0.93-0.96) | 0.88 to 0.99 | 0.22 |
| Operated and non-surgically managed patients, without CA125 | 1 (17) | 0.94 (0.91-0.95) | 0.82 to 0.98 | 0.27 |

Sensitivity and specificity at the 10% risk of malignancy threshold in operated patients were reported in ten studies for ADNEX without CA125 (**Tables S4 and S7**).[22,26,31,37,43,45,46,49,56,61] The summary sensitivity and specificity were 0.93 (95% CI 0.90-0.95, 95% PI 0.73-0.99) and 0.75 (95% CI 0.70-0.79, 95% PI 0.46-0.91), respectively (**Table S8, Figure 6**). For ADNEX with CA125, sensitivity and specificity at the 10% risk of malignancy threshold in operated patients were reported in 24 studies (**Tables S4 and S7)**.[9,18,22,26,30,35–37,40,41,43–46,48,49,51,55,56,58–61,100] The summary sensitivity and specificity were 0.94 (CI 95% 0.92-0.95; 95% PI 0.80-0.98) and 0.77 (CI 95% 0.73-0.81; 95% PI 0.47-0.93, respectively (**Table S8, Figure 7**).

**Figure 6.**
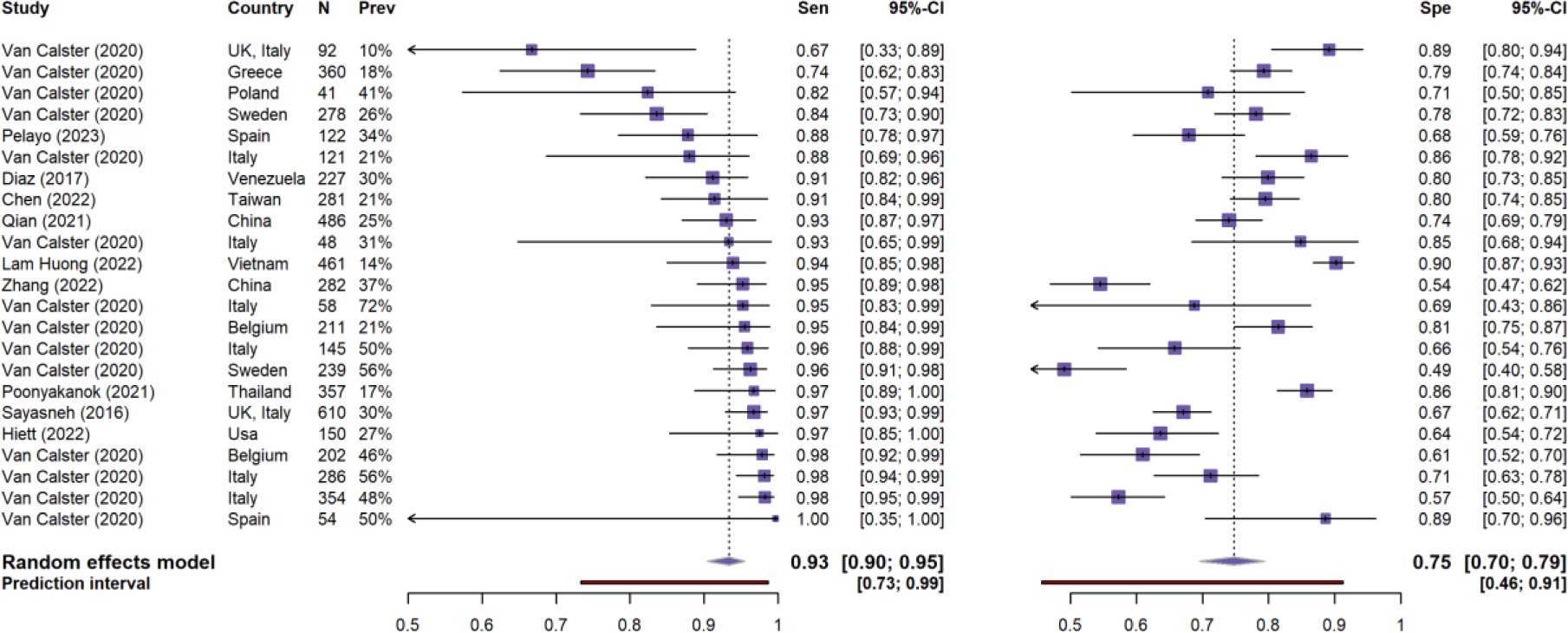
Forest plot of sensitivity (left) and specificity (right) at the 10% risk of malignancy threshold for ADNEX without CA125. CI, confidence interval Prev, prevalence of malignancy; Sen, sensitivity; Spe, specificity.

**Figure 7.**
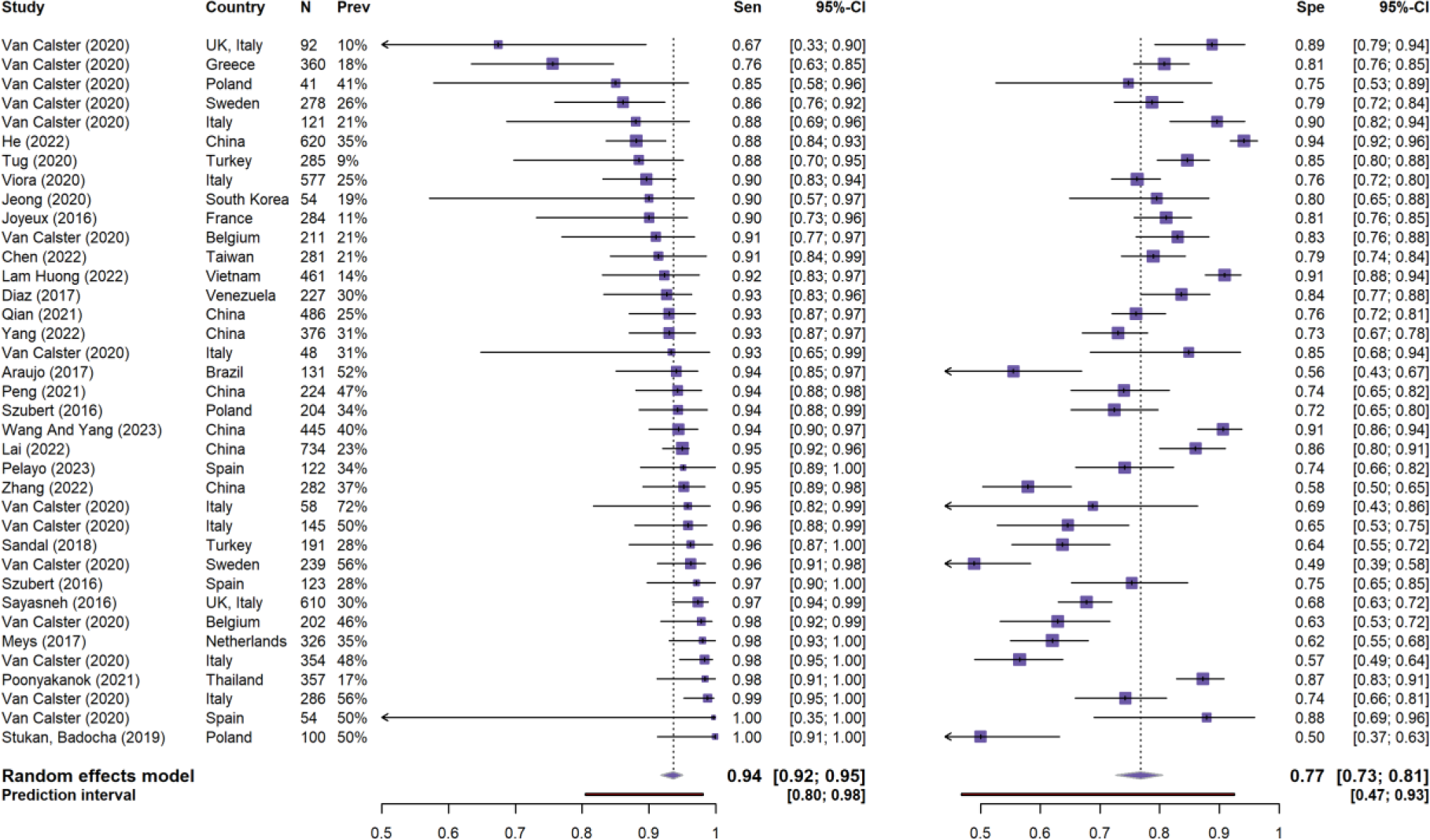
Forest plot of sensitivity (left) and specificity (right) at the 10% risk of malignancy threshold for ADNEX with CA125. CI, confidence interval Prev, prevalence of malignancy; Sen, sensitivity; Spe, specificity.

NB and RU were calculated using studies that presented sensitivity and specificity at the 10% risk of malignancy threshold in operated patients (**Table S7)**. For ADNEX without CA125, the summary NB was 0.28 (95% CrI 0.22-0.35, 95% PI 0.05-0.67) and the summary RU 0.50 (95% CrI 0.36-0.62, 95% PI -0.41-0.81). The probability that the model is clinically useful in a random new centre was estimated to be 91% (**Table S9**, **Figures S6-7**). For ADNEX with CA125, the summary NB was 0.27 (95% CrI 0.23-0.33, 95% PI 0.06-0.64) and the summary RU 0.54 (95% CrI 0.45-0.61, 95% PI -0.12-0.78). The probability that the model is clinically useful in a random new centre was estimated to be 95% (**Table S9, Figures S8-9**).

Pairwise AUCs in operated patients were reported in 4 studies for ADNEX without CA125 and in 5 studies for ADNEX with CA125 (**Table S10**). [58,101–104] The summary pairwise AUCs for ADNEX without CA125 ranged from 0.66 (stage II-IV primary invasive vs metastatic) to 0.97 (benign vs stage II-IV primary invasive) (**Table S11**). For ADNEX with CA125, the summary pairwise AUCs ranged from 0.72 (borderline vs stage I primary invasive) to 0.98 (benign vs stage II-IV primary invasive).

The AUC for benign vs malignant tumours in operated and non-surgically managed patients combined was reported in 2 studies (5167 tumours, 18 centres, 8 countries) with a summary estimate of 0.94 (95% CI 0.93-0.96, 95% PI 0.88-0.99) for ADNEX with CA125 **(Table 2**).[8,104] ADNEX without CA125 was assessed in only 1 study (4905 tumours, 17 centres, 7 countries). [104] This study reported summary AUC 0.94 (95% CI 0.91-0.95, 95% PI 0.82-0.98).

Sensitivity and subgroup results for AUC, specificity, sensitivity, NB and RU are shown in **Tables 2 and S6-9**. These results showed that findings were robust (AUCs ranged from 0.90 to 0.95 across all analyses) and clinical utility was suggested in all subgroups. When limiting the analyses to low RoB studies, summary AUCs were 0.93 (with CA125) and 0.91 (without CA125). Sensitivity was higher and specificity lower in oncology versus non-oncology centres and in postmenopausal versus premenopausal patients. In line with this, meta-regression suggested that prevalence of malignancy was not related to AUC but positively to sensitivity and negatively to specificity (**Figures S10-12**).

Meta-analysis of calibration in only operated patients was not feasible as only one study reported calibration slope and intercept.[104] Four studies presented a calibration plot: in three studies [102,103,105] the estimated risks were close to the observed ones, in one study [104] the risk of malignancy was slightly underestimated.

Based on the subdomains of GRADE assessment, we have identified the risk of bias in the studies included in this meta-analysis as a significant limitation affecting the certainty of our meta-analysis results. Two studies (representing 5511 tumours or 32% of total tumours included in this review) did not fall under the classification of high risk of bias. [103,104] However, the sensitivity analysis according to risk of bias showed consistent findings (Table 2, Table S8 and S9). Funnel plots for the AUC did not suggest publication bias **(Figure S13).**

## DISCUSSION

The ADNEX model exhibited robust discrimination and classification performance between benign and malignant tumours across various settings and populations. In addition, the results indicate that ADNEX has clinical utility at the 10% risk of malignancy threshold, for example to decide whether a patient should be referred for assessment in a gynaecological oncology centre.

We found deficiencies in study reporting, and most studies were judged to be at high risk of bias. However, our sensitivity analyses indicated that performance was almost identical in low risk and high risk of bias studies. For ADNEX with CA125, the AUC was 0.93 based on all studies vs 0.93 when based on only low risk of bias studies. For ADNEX without CA125, the AUCs were 0.93 (all studies) and 0.91 (low risk of bias only). High risk of bias was mainly caused by low sample size, absence of calibration performance, and unjustified use of complete case analysis. Low sample size makes the estimated AUC less precise but does not systematically affect the AUC, unless there is publication bias. The funnel plots do not suggest publication bias. Absence of calibration does not affect the AUC. Using complete cases regarding CA125 or other predictors may lead to underestimation of the AUC, because missing values tend to be associated with the examiner’s subjective impression that the tumour is benign.[106,107] Complete case analysis would then tend to exclude clearly benign tumours, which would make the sample more homogeneous and reduce the AUC. However, our results suggest that the impact of complete case analysis on performance may have been minimal. The impact on calibration could not be assessed. Taken together, the results presented in our meta-analysis are reliable.

Strengths of our systematic review include the meta-analysis of ADNEX both as a risk model and as a diagnostic test, and the thorough critical appraisal regarding risk of bias and reporting quality using recommended checklists.[79,108] Our study also has limitations. First, some study authors have a conflict of interest because they were involved in developing ADNEX or in some of the included external validation studies. To address this, independent researchers with expertise in study methodology and prediction modelling evaluated the IOTA-related studies. Second, calibration performance was reported in only four studies, and meta-analysis of calibration was not possible.

Previous systematic reviews have conducted meta-analyses of the diagnostic performance of ADNEX with CA125.[62–67] They used the QUADAS-2 tool [109] to assess risk of bias, and found between 0 and 64% of studies to be at high risk of bias in at least one domain. We identified 45/47 studies as high risk of bias using the PROBAST tool designed for the appraisal of risk prediction models. None of the previous meta-analyses of ADNEX included clinical utility, calibration or AUC. Our results align with those of other systematic reviews of risk prediction modelling studies in various domains. These consistently indicate that reporting in the original studies was poor and that many studies were at high risk of bias.[110–116]. Still, the results in terms of sensitivity and specificity of ADNEX in our meta-analysis were similar to those in the other meta-analyses of ADNEX performance.

Our findings support the use of ADNEX (1) to decide where, by whom and how to operate when a decision to operate a patient has already been taken (meta-analysis in operated patients), or (2) to choose between surgery and conservative follow-up (meta-analysis in operated and non-surgically managed patients). Because the AUC of ADNEX with and without CA125 are similar, and because adding CA125 mainly helps to distinguish between different types of malignant tumours, we argue that the main use of ADNEX without CA125 is to support the decision whether to operate or follow non-surgically, whereas the main use of ADNEX with CA125 is to support decisions regarding type of surgery. Therefore, we believe that measuring CA125 is often not needed.

While our findings suggest that ADNEX has clinically utility, well conducted validations of any model are always of value to monitor its performance in diverse regions and clinical settings, and over time.[117] In line with this, to improve ADNEX performance even further, efforts to update the ADNEX formula are of interest.[117,118] If further validation studies are conducted, we recommend to include a validation of ADNEX without CA125, to obtain a sufficiently large sample to assess calibration and multinomial discrimination, and to include patients irrespective of whether they are managed surgically or non-surgically, despite the challenges regarding reference standard for non-surgically managed patients.[87,119] Methodological recommendations for validation studies include using available tools to guarantee adequate sample size, describe missing data in detail and use methods such as imputation when needed, and assess calibration performance.[118,120–124] Finally, adherence to the TRIPOD reporting checklists is important to maximise the value of the validation study (www.tripod-statement.org).

## CONCLUSION

ADNEX has been widely validated with AUC values >0.90 to discriminate between benign and malignant tumours across various settings, and with strong results regarding clinical utility at the 10% risk of malignancy threshold.

## Supporting information

PRISMA 2020 Abstract Checklist

PRISMA 2020 Checklist

TRIPOD SRMA Checklist

## Data Availability

The data and code used for the analysis and the generation of the figures is publicly available in Open Science Framework (https://osf.io/jtsvd/). [80] This includes the extraction sheet, one script for running the meta-analysis and one script for generating all the figures included in the paper (except PRISMA Flowchart). Note that for some rows in the extraction sheet we deleted the performance measures for the menopausal subgroups because this information was provided by authors upon request therefore it is not publicly available.

## ACKNOWLEDGMENT

The authors wish to thank Thomas Vandendriessche, Chayenne Van Meel and Krizia Tuand, the biomedical reference librarians of the KU Leuven Libraries – 2Bergen – learning Centre Désiré Collen (Leuven, Belgium), for their help in conducting the systematic literature search.

We gratefully acknowledge the authors of the external validation of ADNEX for providing their valuable data.

This research will be presented at ISUOG World Congress 2023 in Seoul (October 2023).

## OTHER INFORMATION

### Transparency declaration

The lead authors affirm that this manuscript is an honest, accurate, and transparent account of the study being reported; that no important aspects of the study have been omitted; and that any discrepancies from the study as planned (and, if relevant, registered) have been explained.

