## Supplementary for "The ADNEX risk prediction model for ovarian cancer diagnosis: A systematic review and meta-analysis of external validation studies"

**SUPPLEMENTARY MATERIAL**

**Corresponding author**

Ben Van Calster

KU Leuven, Department of Development and Regeneration

Herestraat 49 box 805

3000 Leuven

Belgium

### S1. SEARCH STRATEGY

**Phase 1:**

The following databases were searched for eligible studies:

Search string for **PubMed**: ADNEX [tiab] OR (assessment[tiab] AND "different neoplasias"[tiab] AND "adnexa"[tiab]) AND ("2014/10/18"[Date - Publication] : "3000"[Date - Publication])

Search string for **EMBASE:** ADNEX:ti,ab,kw OR (assessment:ti,ab,kw AND 'different neoplasias':ti,ab,kw AND 'adnexa':ti,ab,kw) AND [2014-2024]/py

Search string for **Web of Science Core Collection**: TS= ("ADNEX" OR ("assessment" AND "different neoplasias" AND "adnexa")) AND PY=(2014-2024)

Search string for **SCOPUS**: TITLE-ABS-KEY (“ADNEX" OR ( "assessment" AND "different neoplasias" AND "adnexa" ) ) AND PUBYEAR > 2013

Search string for **EuropePMC**: adnex AND (SRC:PPR)

Additionally all the citations of ADNEX original paper (https://pubmed.ncbi.nlm.nih.gov/25320247/ ) were retrieved in **PubMed, SCOPUS/EMBASE and Web of Science**. We also searched for studies in systematic reviews that mention the ADNEX model.

**Phase 2:**

For the included articles in phase 1, all the relevant citations and references not already checked in phase 1 were checked in **PubMed, SCOPUS/EMBASE and Web of Science** for inclusion assessment. To determine if a referenced paper or citation was relevant to the systematic review, the title and context in the included paper were used as recommended by Wohlin.[1] Phase 2 was performed at data extraction for included papers in phase 1.

The search was first conducted in 29th November 2022 and repeated in 3rd March 2023 and 15th May 2023.

Two articles were written in a language that was not understood by any of the co-authors (Turkish and Indonesian). Because we did not find suitable translators, we used the automatic translation tool deepl.com.

### S2. IOTA STEERING COMMITTEE

The members of the IOTA steering committee are:

Dirk Timmerman, MD PhD (founder and coordinator, gynaecologist, KU Leuven, Belgium)

Tom Bourne, MD PhD (founder, gynaecologist, Imperial College London, UK)

Lil Valentin, MD PhD (founder, gynaecologist, Lund University, Sweden)

Antonia Testa, MD PhD (gynaecologist, Università Cattolica del Sacro Cuore, Rome, Italy)

Wouter Froyman, MD PhD (gynaecologist, KU Leuven, Belgium)

Ben Van Calster, PhD (medical statistician, KU Leuven, Belgium)

### S3. META-ANALYSIS METHODS FOR AUC, SENSITIVITY, SPECIFICITY

We first extracted all the performance metrics as they were reported in the studies. Some studies presented confidence intervals or standard errors for the metrics, others did not. We used the following methods to approximate the uncertainty.

Approximation of the standard error for the logit of the AUC was based on Newcombe's method 4 [2] implemented in the ccalc function from the "metamisc" R package [3]:

$$SE(logit\ c) \approx \frac{SE(c)}{c(1-c)} \approx \sqrt{\frac{1 + n^* \frac{1-c}{2-c} + \frac{(m^* c)}{1+c}}{mnc(1-c)}},$$

where c is the *c* statistic, *n* the number malignant tumours and *m* the number of benign tumours, and $n^* = m^* = \frac{1}{2}(m+n)$.

Approximation of standard error for sensitivity and specificity was based on Wilson's method [4,5] implemented in madad function from the mada R package [6]:

$$CI = \frac{\hat{p} + \frac{z^2}{2n} \pm z\sqrt{\frac{\hat{p}(1-\hat{p})}{n} + \frac{z^2}{4n^2}}}{1 + \frac{z^2}{n}},$$

with $\hat{p}$ as reported sensitivity/specificity, *n* the number of tumours, and $z$ denotes the quantile of the standard normal distribution.

To obtain the summary estimates, we performed a random effects meta-analysis to account for differences between studies. Random effects weights were calculated with inverse variance method $w_k = \frac{1}{s_k^2 - \tau^2}$ with $s_k^2$ as the within study variance and $\tau^2$ as the between study variance. We calculated $\tau^2$ using Restricted Maximum Likelihood ("REML") [7].

Subgroup analysis was performed by selecting from the k studies the studies that were part of the subgroup and conducting the meta-analysis independently from the whole sample, hence we did not use a common $\tau^2$. To assess the association of prevalence of malignancy with the AUC and sensitivity and specificity at the 10% risk of malignancy threshold, we used random effects meta-regression. This method is similar to a traditional regression but instead of patients as unit of analysis we have studies. Then the model takes into account between study heterogeneity and within study heterogeneity in the same way as meta-analysis. [8]

Confidence intervals of the random effects meta analysis are constructed using Sidik-Jonkman Hartung-Knapp method [9,10]:

$$Var_{HKSJ} = \frac{\sum w_k \left(\theta_i - \hat{\theta}\right)^2}{K - 1 \sum w_k}.$$

Prediction intervals for the AUC were calculated using Bayesian methods. We used weak priors based on half Student-t distribution with location m = 0, scale σ = 0.5 and ν degrees of freedom = 3 [3].

$$\tau_{discr} \sim \; Student(0, 0.5^2, 3)\; T\; [0, 10]$$

Prediction intervals for specificity and sensitivity were calculated under the assumption that the random effects of each study are normally distributed with between study standard deviation ($\tau$) as follows:

$$PI = \hat{\mu} \pm \; t_{k-2}\sqrt{\hat{\tau} + SE(\hat{\mu})^2},$$

with $\hat{\mu}$ as the estimated pooled effect, $t_{k-2}$ is the $100\left(1 - \frac{\alpha}{2}\right)$ percentile of the t-student distribution with k-2 degrees of freedom, and k the number of studies in the meta-analysis.

### S4. TRIVARIATE RANDOM EFFECTS META-ANALYSIS FOR NET BENEFIT

We conducted a random effects meta-analysis of Net Benefit (NB) of ADNEX at the 10% risk of malignancy threshold according to the methodology described in [11]. Relative Utility (RU) expresses NB as a percentage of the maximum possible utility: RU=1 indicates maximum possible utility, RU=0 means no utility, RU<0 means harm. No utility means that NB of ADNEX is not higher than NB of treating all patients ($NB_{TA}$) and NB of treating no one ($NB_{TN}$, which is 0 by definition). In that case, ADNEX is not better than simply assuming that everyone needs treatment or that no one needs treatment without the use of any model.  Harm means that NB of ADNEX is lower than $NB_{TA}$ or $NB_{TN}$. Harm means that you can make better decisions without the model.

NB, $NB_{TA}$, and RU are defined as follows:

$$NB = Se * P - w * (1 - Sp) * (1 - P),$$

$$NB_{TA} = P - w * (1 - P),$$

$$RU = \frac{NB - \max(0, NB_{TA})}{P - \max(0, NB_{TA})},$$

with $Se$ sensitivity, $P$ prevalence of malignancy, $Sp$ specificity, and $w$ the odds of the risk threshold. In our case, $w = 1/9$.

We used weak prior distributions for the sensitivity, specificity and prevalence based on the results reported in the ADNEX model development study (sensitivity 0.965, specificity 0.713, average centre-specific prevalence 0.330). We used normal priors with $Z \sim N(\log\left(\frac{0.965}{1-0.965}\right), \sqrt{1/0.05})$ $Z \sim N(\log\left(\frac{0.713}{1-0.713}\right), \sqrt{(1/0.5)})$ , $Z \sim N(\log\left(\frac{0.330}{1-0.330}\right), \sqrt{(1/0.5)})$ for sensitivity, specificity and prevalence of malignancy, respectively. This assures prior probability distributions bounded by 0 and 1. All other priors were the default priors suggested by Wynants et al [11].

For Markov chain Monte Carlo (MCMC) sampling, we used 1000 samples per chain and a burn-in of 1000, and two chains. This was sufficient for convergence and to have MC error <5% of the standard deviation of the posterior distribution for all parameters of interest.

Note that we implemented the methodology described in (63) after considering a recent erratum [12]

#### S5. PUBLICATION BIAS

Publication bias and small study effects were explored with funnel plots for the AUC. We used random effects model to estimate the summary effect, and tau-squared was estimated using "REML" (Restricted maximum-likelihood estimator). The included studies were the same as those used in the meta-analysis of the AUC with CA125 (see Table S9, row 1).

The funnel plots should be interpreted with caution because deviations from the summary result can be due to publication bias or to differences in case-mix.

### S6. INCONSISTENCIES IN REPORTED DATA

Chen et al (2022) mentioned that there were 281 patients in Table 5 vs 279 in Supplementary Material. We opted for 281 because the calculated 2x2 table for specificity and sensitivity yielded numbers closer to integers.

Joyeux et al (2016) reported results for younger (up to 42 years) versus older (≥43 years) patients in Table 4. The number of patients in the older group was reported to be 188, but the authors sometimes reported that the younger group contained 96 patients and sometimes that it contained 97 patients. As the total sample size is 284, we decided that there were 96 younger patients.

Lam Huong (2022) reported in the abstract that there were 65 malignant and 361 benign tumours, but that the total sample size was 461. In the text and in tables, the authors reported on 396 benign tumours. We assumed that there was a typing error in the abstract and extracted 396 benign cases and 65 malignancies.

Nam et al (2021) reported a sensitivity of 0.90 with 13 malignancies which is not possible. It should be either 0.92 with 12 true positives or 0.84 with 11 true positives. For this reason, we decided to exclude this study from the meta-analysis of specificity and sensitivity.

Tavoraité et al (2021) reported a specificity of 0.46, but with the reported number of true negatives it should have been either 0.48 or 0.45. This study was not eligible for meta-analysis because they analysed ADNEX by the level of expertise of the examiners. According to the reported accuracy, we assumed the correct specificity to be 0.48 with 16 true negatives, 17 true positives, 17 false positives, and 0 false negatives.

Liu et al (2021) reported on sensitivity and specificity, yet only provided information on the number of malignant masses in the study. We decided to exclude this study from all numerical analysis, but we included the 84 malignancies reported by Liu et al in the total number of tumours in our systematic review.

Szubert et al (2020) reported on AUC and sensitivity and specificity for three subgroups that are stratified by the certainty of the subjective assessment: tumours for which the examiner states that it is ‘certainly benign’ or ‘certainly malignant’, tumours for which the examiner states it is ‘probably benign’ or ‘probably malignant’, and tumours for which the examiner is uncertain. However, for the group of certain tumours, the AUC and sensitivity were 1 but the specificity was not perfect. This is not possible, and we could not check the source of the inconsistency, because the authors did not report the number of cases in each subgroup. For this reason, we decided not to include this study’s metrics in our paper. This study was also not included in any meta-analysis, because no performance was reported for all tumours (irrespective of subjective assessment). This study is labelled as one that focuses on specific clinical subgroups, in this case subgroups depending on the subjective assessment of the examiner.

### TABLES

#### Table S1. Data extracted from each validation study.

| | Item | Values |
|---|---|---|
| **General information** | Name of the reviewer | Name |
| | Number of validations (e.g. total study population plus postmenopausal gives two validations) | Number of validations |
| | Unit of study | Patient or tumour |
| | Version of ADNEX | With CA125, without CA125, unclear |
| **Target population and setting** | Single country | Yes or No |
| | Number of countries | Number of countries |
| | Start date and end date of recruitment | Start and end date |
| | Target population | Operated only, or both operated and managed with follow-up |
| **Study description** | Study design | Prospective, retrospective, ambispective, unclear cohort. |
| | Setting | Oncology, Non-oncology, unclear |
| | Recruitment method | Consecutive, probably consecutive, other, unclear |
| | Number of centres | Monocentric or multicentric and number of centres |
| | Inclusion criteria & Exclusion criteria | Listed as in the study |
| | Missing data as exclusion criteria | Yes, No, unclear |
| | Exclusion variables | ADNEX predictors or outcome with missing data that resulted in exclusion of the patient or tumour |
| | Number of excluded patients because of missing data | Number of excluded patients |
| | How are borderline ovarian tumours treated | Benign, malignant, other |
| **Predictors** | Measurement used in the study | Mean, Median, Unclear, Not reported |
| | Variability measure | Standard deviation, Interquartile range, Range, Unclear, Not reported |
| | Age of the population | Age (Variability) |
| | Reported descriptive statistics for the ADNEX predictors | Age; CA125; Family history; Maximal diameter; Solid tissue; Papillary projections; >10 cyst locules; Shadows; Ascites |
| | Reported descriptive statistics of predictors by outcome | Total; Benign; Borderline; Stage I; Stage II-IV; Metastasic; Malignant |
| **Subgroup analysis** | Menopausal data | Yes or No |
| | Conservative follow-up | Yes or No; if yes, duration of follow up |
| **Outcome** | Type | Multinomial or binary |
| | Reference standard | Histology, Other |
| | Sample size and number of malignancies and tumour subtypes | Number |
| | Malignancy rate | Percentage |
| **Analysis** | Missing data | Number of patients with missing information for any variable |
| | Handling of missing data | Complete case analysis, single imputation, multiple imputation, not reported |
| | Software for missing data | Python, R, Stata, SAS, Not reported |
| | AUC as the sum of two triangles | Yes or no |
| | Performance AUC Benign vs Malignant | AUC (CI 95%) |
| | AUC ROC plot | Yes or No |
| | Pairwise AUC methodology | Conditional risk, other, not reported |
| | Performance pairwise AUC | AUC (CI 95%) for the 10 possible pairs |
| | Performance PDI | PDI |
| | Calibration plot | Yes or no |
| | Calibration intercept and slope | Calibration slope and intercept |
| | Multinomial calibration plot | Yes or No |
| | Risk of malignancy cut-off | Cut-off |
| | Sensitivity/Specificity at all cut-offs | Sensitivity, specificity (CI 95%) |
| | PPV and NPV at 10% | PPV, NPV (CI 95%) |
| | DOR at 10% | DOR |
| | Net benefit | Net benefit |
| | Extra reported metrics | Names of metrics |
| **General information** | Statistical software used | Name of software(s) |
| | Conclusion or general opinion on ADNEX | Text |
| **TRIPOD** | All applicable tripod items (See Table S3) | Yes or No, if NO with an explanation for our classifying an item as not having been addressed by the authors |
| **PROBAST** | All applicable signalling questions and risk of bias (ROB) assessment by subdomain and overall ROB | Yes, Probably Yes, No, Probably no, No information for signalling questions.<br>Low, Unclear, High ROB<br>Arguments for ROB classification when needed |

AUC, area under the receiver operating characteristic curve; ROC, receiver operating characteristic curve; PDI, polytomous discrimination index; PPV, positive predictive value; NPV, negative predictive value; DOR, diagnostic odds ratio; CI, confidence interval.

**Table S2. Transparent reporting of a multivariable prediction model for individual prognosis or diagnosis (TRIPOD) items.**

| Section/Topic | Item | Checklist Item |
|---|---|---|
| Title | 1 | Identify the study as developing and/or validating a multivariable prediction model, the target population, and the outcome to be predicted. |
| Abstract | 2 | Provide a summary of objectives, study design, setting, participants, sample size, predictors, outcome, statistical analysis, results, and conclusions. |
| Background and objectives | 3a | Explain the medical context (including whether diagnostic or prognostic) and rationale for developing or validating the multivariable prediction model, including references to existing models. |
| | 3b | Specify the objectives, including whether the study describes the development or validation of the model or both. |
| Source of data | 4a | Describe the study design or source of data (e.g., randomized trial, cohort, or registry data), separately for the development and validation data sets, if applicable. |
| | 4b | Specify the key study dates, including start of accrual; end of accrual; and, if applicable, end of follow-up. |
| Participants | 5a | Specify key elements of the study setting (e.g., primary care, secondary care, general population) including number and location of centres. |
| | 5b | Describe eligibility criteria for participants. |
| | 5c | Give details of treatments received, if relevant. |
| Outcome | 6a | Clearly define the outcome that is predicted by the prediction model, including how and when assessed. |
| | 6b | Report any actions to blind assessment of the outcome to be predicted. |
| Predictors | 7a | Clearly define all predictors used in developing or validating the multivariable prediction model, including how and when they were measured. |
| | 7b | Report any actions to blind assessment of predictors for the outcome and other predictors. |
| Sample size | 8 | Explain how the study size was arrived at. |
| Missing data | 9 | Describe how missing data were handled (e.g., complete-case analysis, single imputation, multiple imputation) with details of any imputation method. |
| Statistical analysis methods | 10c | For validation, describe how the predictions were calculated. |
| | 10d | Specify all measures used to assess model performance and, if relevant, to compare multiple models. |
| Risk groups | 11 | Provide details on how risk groups were created, if done. |
| Development vs. validation | 12 | For validation, identify any differences from the development data in setting, eligibility criteria, outcome, and predictors. |
| Participants | 13a | Describe the flow of participants through the study, including the number of participants with and without the outcome and, if applicable, a summary of the follow-up time. A diagram may be helpful. |
| | 13b | Describe the characteristics of the participants (basic demographics, clinical features, available predictors), including the number of participants with missing data for predictors and outcome. |
| | 13c | For validation, show a comparison with the development data of the distribution of important variables (demographics, predictors and outcome). |
| Model performance | 16 | Report performance measures (with CIs) for the prediction model. |
| Limitations | 18 | Discuss any limitations of the study (such as nonrepresentative sample, few events per predictor, missing data). |
| Interpretation | 19a | For validation, discuss the results with reference to performance in the development data, and any other validation data. |
| | 19b | Give an overall interpretation of the results, considering objectives, limitations, results from similar studies, and other relevant evidence. |
| Implications | 20 | Discuss the potential clinical use of the model and implications for future research. |
| Supplementary information | 21 | Provide information about the availability of supplementary resources, such as study protocol, Web calculator, and data sets. |
| Funding | 22 | Give the source of funding and the role of the funders for the present study. |

For more information see [13,14]

**Table S3. Descriptive characteristics of included studies (n=47).**

| Study | Region | Type of centre | Number of centres | Unit | Clin/Histo Focus[1] | N (benign - malignant) | ADNEX version |
|---|---|---|---|---|---|---|---|
| **Epstein (2016) [15]** [2] | Europe | Oncology | 1 | Patient | Yes | 126 (0-126) | With CA125 |
| **Joyeux (2016) [16]** | Europe | Unclear | 2 | Patient | No | 284 (254-30) | With CA125 |
| **Sayasneh (2016) [17]** [2] | Europe | Oncology | 3 | Patient | No | 610 (428-182) | Both |
| **Szubert (2016) [18]** [2] | Europe | Oncology | 1 | Patient | No | 204 (134-70) | Mixed[3] |
| **Araujo (2017) [19]** | South America | Oncology | 1 | Patient | No | 131 (63-68) | With CA125 |
| **Diaz (2017) [20]** | South America | Oncology | 1 | Patient | No | 227 (159-68) | Both |
| **Meys (2017) [21]** [2] | Europe | Oncology | 1 | Patient | No | 326 (211-115) | With CA125 |
| **Sandal (2018) [22]** | Asia | Oncology | 1 | Patient | No | 191 (138-53) | With CA125 |
| **Chen (2019) [23]** | Asia | Oncology | 1 | Patient | No | 278 (203-75) | Both |
| **Nohuz (2019) [24]** | Europe | Oncology | 1 | Patient | Yes | 93 (89-4) | With CA125 |
| **Stukan, Alcazar (2019) [25]** [2] | Europe | Oncology | 7 | Patient | Yes | 162 (0-162) | Both |
| **Stukan, Badocha (2019) [26]** | Europe | Oncology | 1 | Patient | No | 100 (50-50) | With CA125 |
| **Gaurilcikas (2020) [27]** | Europe | Oncology | 1 | Patient | Yes | 85 (0-85) | Both |
| **Jeong (2020) [28]** | Asia | Oncology | 1 | Patient | No | 59 (49-10) | With CA125 |
| **Quaranta (2020) [29]** | Europe | Oncology | 1 | Patient | Yes | 34 (34-0) | With CA125 |
| **Szubert (2020) [30]** | Europe | Oncology | 2 | Tumor | Yes | 451 (250-201) | With CA125 |
| **Tug (2020) [31]** | Asia | Unclear | 1 | Patient | No | 285 (259-26) | With CA125 |
| **Van Calster (2020) [32]** [2] | Europe | Both | 17 | Patient | No | 4905 (3864-1041) | Both |
| **Viora (2020) [33]** | Europe | Non-oncology | 1 | Patient | No | 577 (433-144) | With CA125 |
| **Butureanu (2021) [34]** | Europe | Unclear | 1 | Patient | No | 230 (223-7) | With CA125 |
| **Czekierdowski (2021) [35]** | Europe | Both | 4 | Patient | Yes | 36 (27-9) | Without CA125 |
| **Jiang (2021) [36]** | Asia | Oncology | 1 | Patient | Yes | 63 (42-21) | With CA125 |
| **Lee (2021) [37]** | Asia | Oncology | 11 | Patient | Yes | 236 (223-13) | Unclear |
| **Liu (2021) [38]** | Asia | Unclear | 1 | Patient | No | Unclear (Unclear-84) | Unclear |
| **Nam (2021) [39]** | Asia | Oncology | 1 | Patient | No | 353 (340-13) | With CA125 |
| **Peng (2021) [40]** | Asia | Oncology | 1 | Patient | No | 224 (119-105) | Both |
| **Poonyakanok (2021) [41]** | Asia | Oncology | 1 | Patient | No | 357 (296-61) | Both |
| **Qian (2021) [42]** | Asia | Oncology | 1 | Patient | No | 486 (366-120) | Both |
| **Tavoraitè (2021) [43]** | Europe | Oncology | 1 | Patient | No | 50 (33-17) | Mixed[3] |
| **Behnamfar (2022) [44]** | Asia | Oncology | 2 | Tumor | No | 284 (260-24) | With CA125 |
| **Budiana (2022) [45]** | Asia | Oncology | 1 | Tumor | No | 88 (38-50) | Unclear |
| **Chen (2022) [46]** | Asia | Oncology | 1 | Patient | No | 322 (264-58) | Both |
| **Esquivel Villabona (2022) [47]** | South America | Oncology | 1 | Tumor | No | 606 (545-61) | Unclear |
| **Hack (2022) [48]** | North America | Oncology | 1 | Tumor | No | 262 (187-75) | Mixed[3] |
| **He (2021) [49]** | Asia | Oncology | 1 | Patient | No | 620 (402-218) | Both |
| **Hiett (2022) [50]** | North America | Oncology | 1 | Patient | No | 150 (110-40) | Without CA125 |
| **Jianhong (2022) [51]** | Asia | Oncology | 1 | Patient | Yes | 23 (15-8) | Both |
| **Lai (2022) [52]** | Asia | Unclear | 1 | Patient | No | 734 (564-170) | With CA125 |
| **Lam Huong (2022) [53]** | Asia | Oncology | 2 | Patient | No | 461 (396-65) | Both |
| **Velayo (2022) [54]** | Asia | Oncology | 2 | Patient | No | 260 (141-119) | With CA125 |
| **Yang (2022) [55]** | Asia | Oncology | 1 | Patient | No | 376 (259-117) | With CA125 |
| **Zhang (2022) [56]** | Asia | Oncology | 1 | Patient | No | 282 (178-104) | Both |
| **Czekierdowski (2023) [57]** | Europe | Both | 3 | Patient | Yes | 108 (62-46) | With CA125 |
| **Hu (2023) [58]** | Asia | Oncology | 2 | Patient | No | 529 (370-159) | Without CA125 |
| **Pelayo (2023) [59]** | Europe | Oncology | 1 | Patient | No | 122 (81-41) | Both |
| **Rashmi (2023) [60]** | Asia | Oncology | 1 | Patient | No | 90 (80-10) | Both |
| **Wang And Yang (2023) [61]** | Asia | Unclear | 1 | Patient | No | 445 (265-180) | With CA125 |

[1]A study was considered to have clinical or histological focus if the study sample consisted of selected histologies (e.g. only borderline tumours), or a selected subgroup of patients (e.g. only pregnant patients).

[2]Papers extracted by authors PD and GSC.

[3]Mixed" means that the authors used ADNEX with CA125 for patients with CA125 data and ADNEX without CA125 for patients without CA125 data.

#### Table S4. Reported performance for distinguishing benign from malignant tumours (63 validations)

| Study | ADNEX With CA125 | Missing Data handling | AUC Benign versus Malignant (95%CI) | Sensitivity at 10% cut-off (95% CI) | Specificity at 10% cut-off (95% CI) | Calibr. | RoB | TRIPOD items reported |
|---|---|---|---|---|---|---|---|---|
| **Epstein (2016)** | Yes | CCA | NR | NR | NR | No | High | 61% |
| **Joyeux (2016)[a,b]** | Yes | CCA | 0.938 (0.899-0.977) | 90 | 81.1 | No | High | 64% |
| **Sayasneh (2016)[a,b]** | Yes | MI/SI | 0.937 (0.915-0.954) | 97.3 (93.5-98.9) | 67.7 (63.0-72.0) | Yes | Low | 86% |
| **Sayasneh (2016)[a,b]** | No | SI | 0.925 (0.902-0.943) | 96.7 (92.9-98.5) | 67.1 (62.5-71.3) | Yes | Low | 86% |
| **Szubert (2016)[a,b]** | Mixed | CCA | 0.907 (0.858 - 0.948) | 94.3 (88.5 - 98.7) | 72.4 (65.1 - 79.7) | No | High | 54% |
| **Araujo (2017)[a,b]** | Yes | CCA | 0.92 (0.88-0.97) | 94.1 | 55.5 | No | High | 68% |
| **Diaz (2017)[a,b]** | Yes | CCA | 0.933 (0.901-0.964) | 92.64 | 83.64 | No | High | 61% |
| **Diaz (2017)[a,b]** | No | CCA | 0.925 (0.892-0.958) | 91.17 | 79.87 | No | High | 64% |
| **Meys (2017) [b]** | Yes | MI | 0.93 (0.89-0.95) | 98 (93-100) | 62 (55-68) | No | High | 64% |
| **Sandal (2018)** | Yes | CCA | NR | 96.2 (87 - 99.5) | 63.7 (55.2- 71.7) | No | High | 50% |
| **Chen (2019)** | Yes | CCA | 0.94 (0.91-0.97) | 93.3 (85-98) | 77.8 (72-83) | No | High | 68% |
| **Chen (2019)** | No | CCA | 0.93 (0.90-0.96) | NR | NR | No | High | 68% |
| **Nohuz (2019)** | Yes | CCA | NR | 100 | 98.8 | No | High | 43% |
| **Stukan, Alcazar (2019)** | Yes | CCA | NR | NR | NR | No | High | 61% |
| **Stukan, Alcazar (2019)** | No | CCA | NR | NR | NR | No | High | 61% |
| **Stukan, Badocha (2019)[a,b]** | Yes | CCA | 0.972 (0.946-0.999) | 100 | 50 | No | High | 75% |
| **Gaurilcikas (2020)** | Yes | CCA | NR | NR | NR | No | High | 54% |
| **Gaurilcikas (2020)** | No | CCA | NR | NR | NR | No | High | 54% |
| **Jeong (2020)[b,c]** | Yes | NR | 0.924 (0.786-1.0) | 90 | 81.6 | No | High | 64% |
| **Quaranta (2020)** | Yes | CCA | NR | NR | NR | No | High | 54% |
| **Szubert (2020)** | Yes | CCA | NR | NR | NR | No | High | 54% |
| **Tug (2020)[a,b]** | Yes | CCA | 0.941 (0.042 ) | 88.5 | 84.6 | No | High | 64% |
| **Van Calster (2020)[a,b]** | Yes | MI | 0.94 (0.92-0.96) | 91.2 (84.8-95.1) | 85.3 (80.9-88.8) | Yes | Low | 100% |
| **Van Calster (2020)[a,b]** | No | MI | 0.94 (0.91-0.95) | 91.1 (84.5-95.1) | 84.5 (80.1-88.0) | Yes | Low | 100% |
| **Viora (2020)[a,b]** | Yes | CCA | 0.9111 (0.8788-0.9389) | 89.6 (83.1-94.0) | 76.2 (71.9-80.1) | Yes | High | 64% |
| **Butureanu (2021)** | Yes | NR | NR | NR | NR | No | High | 43% |
| **Czekierdowski (2021)** | No | CCA | NR | NR | NR | No | High | 64% |
| **Jiang (2021)** | Yes | NR | NR | NR | NR | No | High | 43% |
| **Lee (2021)** | Unc | NR | 0.709 (0.646-0.766) | NR | NR | No | High | 50% |
| **Liu (2021) [c]** | Unc | NR | 0.821 | NR | NR | No | High | 39% |
| **Nam (2021)[a,b]** | Yes | NR | 0.92 | 90 | 82.0 | No | High | 61% |
| **Peng (2021)[a]** | Yes | NR | 0.94 (0.90-0.97) | 94.3 (88.0-97.9) | 74.0 (65.1-81.6) | No | High | 64% |
| **Peng (2021)[a,b]** | No | NR | 0.93 (0.89-0.96) | NR | NR | No | High | 68% |
| **Poonyakanok (2021)[a,b]** | Yes | CCA | 0.975 (0.953-0.988) | 98.4 (91.2-100) | 87.2 (82.8-90.8) | No | High | 75% |
| **Poonyakanok (2021)[a,b]** | No | CCA | 0.972 (0.949-0.987) | 96.7 (88.7-99.6) | 85.8 (81.3-89.6) | No | High | 79% |
| **Qian (2021)[a,b]** | Yes | CCA | 0.94 (0.92-0.96) | 93 (87-97) | 76 (72-81) | No | High | 68% |
| **Qian (2021)[a,b]** | No | CCA | 0.94 (0.91-0.96) | 93 (87-97) | 74 (69-79) | No | High | 68% |
| **Tavoraitė (2021)** | Mixed | CCA | NR | 100 (80.5-100) | 81.8 (64.5-93) | No | High | 46% |
| **Behnamfar (2022)[a]** | Yes | NR | 0.746 (0.691-0.796) | NR | NR | No | High | 46% |
| **Budiana (2022)** | Unc | NR | NR | NR | NR | No | High | 46% |
| **Chen (2022)[a]** | Yes | CCA | 0.95 (0.91-0.97) | 91.4 (84.2-98.6) | 78.9 (73.6-84.3) | No | High | 75% |
| **Chen (2022)[a]** | No | CCA | 0.94 (0.91-0.97) | 91.4 (84.2-98.6) | 79.5 (74.2-84.6) | No | High | 75% |
| **Esquivel Villabona (2022)[c]** | Unc | NR | 0.895 | 91.8 (82.2-96.4) | 87.2 (84.08-89.71) | No | High | 64% |
| **Hack (2022)** | Mixed | CCA | 0.9479 | NR | NR | No | High | 61% |
| **He (2022)[a]** | Yes | CCA | 0.97 (0.96-0.98) | 88.06 (83.58-92.54) | 94.10 (91.79-96.41) | No | High | 71% |
| **He (2022)[a,b]** | No | CCA | 0.97 (0.95-0.98) | NR | NR | No | High | 71% |
| **Hiett (2022)[a,b]** | No | NR | 0.937 | 97.5 (85.3-99.9) | 63.6 (53.9-72.4) | No | High | 61% |
| **Jianhong (2022)** | Yes | CCA | 0.892 (0.692-0.982) | 87.50 (47.3-99.7) | 73.33 (44.9-92.2) | No | High | 57% |
| **Jianhong (2022)** | No | CCA | 0.896 (0.697-0.983) | 100.0 (63.1-100.0) | 53.3 (26.6-78.7) | No | High | 57% |
| **Lai (2022)[bc]** | Yes | CCA | 0.90 (0.87-0.94) | 95 (92-96) | 86 (80-91) | No | High | 57% |
| **Lam Huong (2022)[a,b]** | Yes | NR | 0.961 (0.939 - 0.977) | 92.3 (83.0 - 97.5) | 90.9 (87.6 - 93.6) | No | High | 43% |
| **Lam Huong (2022)[a,b]** | No | NR | 0.956 (0.933 - 0.973) | 93.9 (85.0 - 98.3) | 90.2 (86.8 - 92.9) | No | High | 46% |
| **Velayo (2022) [c]** | Yes | CCA | 0.78 (0.73-0.82) | NR | NR | No | High | 54% |
| **Yang (2022)[a,b]** | Yes | CCA | 0.914 (0.881-0.941) | 93 (87-97) | 73 (67-78) | No | High | 71% |
| **Zhang (2022)[a,b]** | Yes | SI | 0.93 (0.89-0.96) | 95.2 (89.1-98.4) | 57.9 (50.3-65.2) | Yes | Unc | 89% |
| **Zhang (2022)[a,b]** | No | SI | 0.91 (0.87-0.94) | 95.2 (89.1-98.4) | 54.5 (46.9-62.0) | Yes | Unc | 89% |
| **Czekierdowski (2023)** | Yes | NR | NR | NR | NR | No | High | 54% |
| **Hu (2023)** | No | CCA | NR | NR | NR | No | High | 61% |
| **Pelayo (2023) [a,b]** | Yes | CCA | 0.88 | 95.1 (88.7-100) | 74.1 (65.9-82.3) | No | High | 61% |
| **Pelayo (2023) [a,b]** | No | CCA | 0.84 | 87.8 (78.4-97.2) | 67.9 (59.5-76.3) | No | High | 61% |
| **Rashmi (2023) [c]** | Yes | No missings | 0.8 | NR | NR | No | High | 57% |
| **Rashmi (2023) [c]** | No | No missings | 0.786 | NR | NR | No | High | 57% |
| **Wang And Yang (2023) [b]** | Yes | CCA | 0.925 (0.897-0.948) | 94.4 (90.0-97.3) | 90.6 (86.4-93.8) | No | High | 50% |

NR, not reported; Unc, unclear; CCA, complete case analysis; SI, single imputation; MI, multiple imputation; Calibr, calibration; ADNEX, Assessment of Different NEoplasias in the adneXa; AUC, Area under the receiver operating characteristic curve; RoB, Risk of bias; TRIPOD, Transparent reporting of a multivariable prediction model for individual prognosis or diagnosis

[a,b] Included in meta-analysis for AUC (a) and/or sensitivity and specificity (b) in operated patients.

[c] Not included in meta-analysis for AUC because they presented an AUC after dichotomising or categorising risks, i.e. the ROC curve has only 1 or a few points. This was either clear from the ROC curve that was shown, or from the fact that the AUC equals the average of sensitivity and specificity.

**Table S5. Reported performance metrics or graphs.**

| Metric/graph | All 47 studies, n (%) | 36 studies without histological or clinical focus, n (%) |
|---|---|---|
| Discrimination benign vs malignant (any) | 34 (72) | 31 (86) |
| AUC benign vs malignant | 34 (72)[a] | 31 (86)[a] |
| Receiver operating characteristic curve shown | 31 (66) | 27 (75) |
| Classification benign vs malignant (any) | 41 (87) | 34 (94) |
| Sensitivity and specificity for malignancy at the 10% cut-off | 31 (66) | 28 (50) |
| Sensitivity and specificity for malignancy at other cut-offs | 29 (62) | 24 (67) |
| Positive and negative predictive value at the 10% cut-off | 22 (47) | 21 (58) |
| Positive and negative predictive value at any cut-offs | 15 (32) | 14 (39) |
| Positive and negative likelihood ratio at any cut-offs | 18 (36) | 15 (42) |
| Diagnostics odds ratio (DOR) at the 10% cut-off | 10 (21) | 10 (28) |
| Accuracy | 8 (17) | 6 (17) |
| Multinomial discrimination (any) | 12 (26) | 12 (33) |
| Pairwise AUC | 12 (26)[b] | 12 (33)[b] |
| Polytomous Discrimination Index (PDI) | 3 (6) | 3 (8) |
| Calibration (any) | 4 (9) | 4 (11) |
| Calibration plot for risk of malignancy | 4 (9) | 4 (11) |
| Calibration intercept and slope for risk of malignancy | 1 (2) | 1 (3) |
| Multinomial calibration plots | 1 (2) | 1 (3) |
| Clinical utility (any) | 1 (2) | 1 (3) |
| Net benefit | 1 (2) | 1 (3) |
| Decision curve (Net benefit over different thresholds) | 1 (2) | 1 (3) |

AUC; Area under the receiver operating characteristic curve.
[a] Four of these reported the AUC after dichotomising or categorising risk (See Table S4 for details).
[b] Four of these reported the methodology for calculating pairwise AUCs.

**Table S6. Descriptive data for the meta-analysis of area under the receiver operating characteristic curve (AUC) for benign vs malignant tumours.**

| Meta-analysis | Studies | Centres | Countries | Patients | TRIPOD adherence | References |
|---|---|---|---|---|---|---|
| *Main analysis* | | | | | | |
| Operated patients, with CA125[a] | 21 | 43 | 18 | 9202 | 67% | [16–21,26,31–33,40–42,44,46,49,53,55,59,62,63] |
| Operated patients, without CA125 | 12 | 31 | 13 | 6309 | 71% | [17,20,32,40–42,46,49,50,53,56,59] |
| *Sensitivity analyses (operated patients)* | | | | | | |
| High/unclear RoB studies, with CA125 | 19 | 23 | 14 | 6103 | 65% | [16,18–21,26,31,33,40–42,44,46,49,53,55,56,59,62] |
| Low RoB studies, with CA125 | 2 | 20 | 7 | 3099 | 88% | [17,32] |
| IOTA studies, with CA125 | 4 | 23 | 8 | 3752 | 73% | [17,21,32,64] |
| Non-IOTA studies, with CA125 | 17 | 20 | 13 | 5450 | 66% | [16,19,20,26,31,33,40–42,44,46,49,53,55,59,62,63] |
| High/unclear RoB studies, without CA125 | 10 | 11 | 7 | 3210 | 68% | [20,40–42,46,49,50,53,56,59] |
| Low RoB studies, without CA125 | 2 | 20 | 7 | 3099 | 88% | [17,32] |
| IOTA studies, without CA125 | 2 | 20 | 7 | 3099 | 88% | [17,32] |
| Non-IOTA studies, without CA125 | 10 | 11 | 7 | 3210 | 68% | [20,40–42,46,49,50,53,56,59] |
| *Subgroup analyses (operated patients)* | | | | | | |
| Asian centres, with CA125 | 11 | 13 | 7 | 4009 | 66% | [31,40–42,44,49,53,55,55,56,62] |
| Asian centres, without CA125 | 7 | 8 | 4 | 2711 | 71% | [40–42,46,49,53,56] |
| Chinese centres, with CA125 | 5 | 5 | 1 | 1988 | 73% | [65–69] |
| Chinese centres, without CA125 | 4 | 4 | 1 | 1612 | 74% | [65–68] |
| European centres, with CA125 | 8 | 28 | 9 | 4835 | 70% | [16,17,21,26,32,33,59] |
| European centres, without CA125 | 3 | 21 | 7 | 3221 | 79% | [17,32,59] |
| Non-oncology centres, with CA125 | 2 | 9 | 4 | 1327 | 82% | [32,33] |
| Non-oncology centres, without CA125 | 1 | 8 | 4 | 750 | 100% | [32] |
| Oncology centres, with CA125 | 18 | 31 | 16 | 7306 | 68% | [19,44,46,20,49,53,62,40–42,18,31,55,17,32,63,26,33,59] |
| Oncology centres, without CA125 | 12 | 23 | 13 | 5559 | 71% | [20,32,40–42,46,49,50,53,56,59] |
| Postmenopausal, with CA125 | 11 | 32 | 14 | 2359 | 69% | [16,17,19–21,26,26,31,32,41,44] |
| Postmenopausal, without CA125 | 4 | 22 | 9 | 1623 | 79% | [17,20,32,41] |
| Premenopausal, with CA125 | 11 | 32 | 14 | 3061 | 69% | [16,17,19–21,26,26,31,32,41,44] |
| Premenopausal, without CA125 | 4 | 22 | 9 | 2060 | 79% | [17,20,32,41] |
| *Target population* | | | | | | |
| Operated and non-surgically managed patients, with CA125 | 2 | 18 | 8 | 5167 | 80% | [32,48] |
| Operated and non-surgically managed patients, without CA125[b] | 1 | 17 | 7 | 4905 | 100% | [32] |

[a] Szubert (2016) indicated the use of ADNEX version without CA125 in 9 patients where the variable was missing but the results were pooled in the ADNEX with CA125 meta-analysis. [b] Results from [32].

AUC, area under the receiver operating characteristic curve; RoB, risk of bias; TRIPOD, Transparent reporting of a multivariable prediction model for individual prognosis or diagnosis

**Table S7. Descriptive data for the meta-analyses of sensitivity and specificity and clinical utility at the 10% risk of malignancy threshold in operated patients.**

| Meta-analysis | Studies | Centres | Countries | Patients | TRIPOD adherence | References |
|---|---|---|---|---|---|---|
| *Main analysis* | | | | | | |
| Operated patients, with CA125 [a] | 23 | 44 | 17 | 9989 | 67% | [16–20,22,26,28,31–33,40–42,46,49,52,53,55,56,59,61] |
| Operated patients, without CA125 | 10 | 29 | 13 | 5465 | 73% | [17,20,32,41,42,46,50,53,56,59] |
| *Sensitivity analyses* | | | | | | |
| High/unclear RoB studies, with CA125 | 21 | 24 | 13 | 6890 | 64% | [16,18–22,26,28,31,33,40–42,46,49,52,53,55,56,59,61] |
| Low RoB studies, with CA125 | 2 | 20 | 7 | 3099 | 93% | [17,32] |
| IOTA studies, with CA125 | 4 | 23 | 8 | 3752 | 76% | [17,18,21,32] |
| Non-IOTA studies, with CA125 | 19 | 21 | 12 | 6237 | 65% | [16,19,20,22,26,28,31,33,40–42,46,49,52,53,55,56,59,61] |
| High/unclear RoB studies, without CA125 | 8 | 9 | 7 | 2366 | 68% | [20,41,42,46,50,53,56] |
| Low RoB studies, without CA125 | 3 | 21 | 8 | 3381 | 92% | [17,32,56] |
| IOTA studies, without CA125 | 2 | 20 | 7 | 3099 | 93% | [17,32] |
| Non-IOTA studies, without CA125 | 8 | 9 | 7 | 2366 | 68% | [20,41,42,46,50,53,56,59] |
| *Subgroup analyses* | | | | | | |
| Asian centres, with CA125 | 13 | 14 | 6 | 4796 | 65% | [22,28,31,40–42,46,49,52,53,55,56,61] |
| Asian centres, without CA125 | 5 | 6 | 4 | 1867 | 71% | [41,42,46,53,56] |
| Chinese centres, with CA125 | 7 | 7 | 1 | 3167 | 67% | [65–71] |
| European centres, with CA125 | 8 | 28 | 9 | 4835 | 71% | [16,17,21,32,33,59,64] |
| European centres, without CA125 | 3 | 21 | 7 | 3221 | 82% | [17,32,59] |
| Non-oncology centres, with CA125 | 2 | 9 | 4 | 1327 | 82% | [32,33] |
| Non-oncology centres, without CA125 | 1 | 8 | 4 | 750 | 100% | [32] |
| Oncology centres, with CA125 | 18 | 30 | 16 | 6914 | 69% | [19–22,28,32,40–42,46,49,53,59,61] |
| Oncology centres, without CA125 | 10 | 21 | 13 | 4715 | 73% | [17,20,32,41,42,46,50,53,56,59] |
| Postmenopausal, with CA125 | 10 | 28 | 14 | 2240 | 68% | [16,19–21,26,26,31,32,41,61] |
| Postmenopausal, without CA125 | 4 | 20 | 10 | 1431 | 77% | [20,32,41,50] |
| Premenopausal, with CA125 | 10 | 28 | 14 | 2731 | 68% | [16,19–21,26,31,32,41,61,64] |
| Premenopausal, without CA125 | 4 | 20 | 10 | 1792 | 76% | [20,32,41,50] |

[a] Szubert (2016) indicated the use of ADNEX version without CA125 in 9 patients where the variable was missing, but we pooled the results in the ADNEX with CA125 meta-analysis

RoB, risk of bias; TRIPOD, Transparent reporting of a multivariable prediction model for individual prognosis or diagnosis

**Table S8. Meta-analysis results of sensitivity and specificity at the 10% risk of malignancy threshold in operated patients.**

| | Sensitivity | | | Specificity | | |
|---|---|---|---|---|---|---|
| **Meta-analysis** | **Summary estimate (95% CI)[a]** | **95% PI[b]** | $\tau^2$ | **Summary estimate (95% CI)[a]** | **95% PI[b]** | $\tau^2$ |
| *Main analysis* | | | | | | |
| Operated patients, with CA125 | 0.94 (0.92-0.95) | 0.80 - 0.98 | 0.37 | 0.77 (0.73-0.81) | 0.47 - 0.93 | 0.41 |
| Operated patients, without CA125 | 0.93 (0.90-0.95) | 0.73 - 0.99 | 0.58 | 0.75 (0.70-0.79) | 0.46 - 0.91 | 0.35 |
| *Sensitivity analysis* | | | | | | |
| High/unclear RoB studies, with CA125 | 0.93 (0.92-0.95) | 0.88 - 0.96 | 0.07 | 0.78 (0.72-0.82) | 0.46 - 0.94 | 0.45 |
| Low RoB studies, with CA125 | 0.94 (0.89-0.96) | 0.63 - 0.99 | 0.94 | 0.75 (0.68-0.81) | 0.43 - 0.92 | 0.37 |
| IOTA studies, with CA125 | 0.94 (0.90-0.96) | 0.69 - 0.99 | 0.82 | 0.74 (0.68-0.79) | 0.46 - 0.90 | 0.30 |
| Non-IOTA studies, with CA125 | 0.93 (0.91-0.94) | 0.88 - 0.96 | 0.07 | 0.79 (0.73-0.84) | 0.45 - 0.94 | 0.48 |
| High/unclear RoB studies, without CA125 | 0.93 (0.90-0.95) | 0.89 - 0.96 | <0.01 | 0.76 (0.67-0.84) | 0.37 - 0.94 | 0.42 |
| Low RoB studies, without CA125 | 0.94 (0.90-0.96) | 0.65 - 0.99 | 0.88 | 0.73 (0.66-0.79) | 0.42 - 0.91 | 0.35 |
| IOTA studies, without CA125 | 0.94 (0.89-0.96) | 0.61 - 0.99 | 0.96 | 0.74 (0.67-0.80) | 0.44 - 0.91 | 0.33 |
| Non-IOTA studies, without CA125 | 0.93 (0.90-0.95) | 0.89 - 0.96 | <0.01 | 0.76 (0.67-0.84) | 0.37 - 0.94 | 0.42 |
| *Subgroup analyses* | | | | | | |
| Asian centres, with CA125 | 0.93 (0.91-0.95) | 0.88 - 0.96 | 0.07 | 0.82 (0.75-0.87) | 0.48 - 0.96 | 0.47 |
| Asian centres, without CA125 | 0.94 (0.90-0.96) | 0.88 - 0.97 | <0.01 | 0.79 (0.65-0.88) | 0.21 - 0.98 | 0.58 |
| Chinese centres, with CA125 | 0.93 (0.91-0.95) | 0.85 - 0.97 | 0.09 | 0.81 (0.70-0.89) | 0.31 - 0.98 | 0.67 |
| European centres, with CA125 | 0.95 (0.91-0.97) | 0.67 - 0.99 | 0.97 | 0.74 (0.68-0.78) | 0.47 - 0.90 | 0.29 |
| European centres, without CA125 | 0.93 (0.89-0.96) | 0.62 - 0.99 | 0.90 | 0.73 (0.67-0.79) | 0.45 - 0.90 | 0.30 |
| Non-oncology centres, with CA125 | 0.88 (0.83-0.91) | 0.81 - 0.92 | <0.01 | 0.83 (0.77-0.87) | 0.64 - 0.93 | 0.10 |
| Non-oncology centres, without CA125 | 0.87 (0.77-0.93) | 0.52 - 0.97 | 0.20 | 0.83 (0.78-0.87) | 0.67 - 0.92 | 0.05 |
| Oncology centres, with CA125 | 0.95 (0.93-0.96) | 0.81 - 0.99 | 0.48 | 0.73 (0.68-0.78) | 0.41 - 0.91 | 0.41 |
| Oncology centres, without CA125 | 0.94 (0.92-0.96) | 0.77 - 0.99 | 0.53 | 0.72 (0.66-0.77) | 0.42 - 0.90 | 0.33 |
| Postmenopausal, with CA125 | 0.97 (0.94-0.98) | 0.69 - >.99 | 1.48 | 0.65 (0.58-0.71) | 0.36 - 0.86 | 0.31 |
| Postmenopausal, without CA125 | 0.96 (0.92-0.98) | 0.59 - >.99 | 1.69 | 0.62 (0.54-0.68) | 0.36 - 0.82 | 0.21 |
| Premenopausal, with CA125 | 0.93 (0.87-0.96) | 0.41 - >.99 | 1.90 | 0.81 (0.75-0.86) | 0.47 - 0.95 | 0.56 |
| Premenopausal, without CA125 | 0.90 (0.84-0.94) | 0.64 - 0.98 | 0.50 | 0.82 (0.76-0.86) | 0.55 - 0.94 | 0.34 |

CI, confidence interval; PI, prediction interval

[a] CI estimated using Wilson's interval **[4]** when not reported

[b] PI calculated as in [72].

#### Table S9. Meta-analysis results of clinical utility at the 10% cut-off in operated patients.

| Meta-analysis | Net Benefit | | Relative utility | | |
|---|---|---|---|---|---|
| | Summary estimate (95% CrI)[a] | 95% PI[b] | Summary estimate (95% CrI)[a] | 95% PI[b] | P(useful)[b] |
| *Main analysis* | | | | | |
| Operated patients, with CA125 | 0.27 (0.23-0.33) | 0.06 - 0.64 | 0.54 (0.45-0.61) | -0.12 - 0.78 | 95% |
| Operated patients, without CA125 | 0.28 (0.22-0.35) | 0.05 - 0.67 | 0.50 (0.36-0.62) | -0.41 - 0.81 | 91% |
| *Sensitivity analyses* | | | | | |
| High/unclear RoB studies, with CA125 | 0.25 (0.19-0.31) | 0.06 - 0.58 | 0.57 (0.46-0.67) | -0.19 - 0.83 | 95% |
| Low RoB studies, with CA125 | 0.32 (0.22-0.43) | 0.04 - 0.75 | 0.45 (0.25-0.61) | -0.66 - 0.78 | 89% |
| IOTA studies, with CA125 | 0.32 (0.23-0.41) | 0.06 - 0.70 | 0.48 (0.33-0.60) | -0.60 - 0.77 | 91% |
| Non-IOTA studies, with CA125 | 0.24 (0.17-0.31) | 0.05 - 0.60 | 0.57 (0.44-0.67) | -0.35 - 0.83 | 93% |
| High/unclear RoB studies, without CA125 | 0.21 (0.15-0.28) | 0.07 - 0.45 | 0.57 (0.33-0.73) | -0.71 - 0.86 | 90% |
| Low RoB studies, without CA125 | 0.32 (0.23-0.43) | 0.04 - 0.72 | 0.43 (0.24-0.57) | -0.76 - 0.77 | 87% |
| IOTA studies, without CA125 | 0.32 (0.23-0.42) | 0.04 - 0.74 | 0.44 (0.24-0.59) | -1.00 - 0.77 | 85% |
| Non-IOTA studies, without CA125 | 0.21 (0.15-0.30) | 0.07 - 0.48 | 0.57 (0.34-0.72) | -0.47 - 0.85 | 91% |
| *Subgroup analyses* | | | | | |
| Asian centres, with CA125 | 0.22 (0.15-0.30) | 0.04 - 0.57 | 0.62 (0.47-0.73) | -0.49 - 0.86 | 92% |
| Asian centres, without CA125 | 0.19 (0.11-0.30) | 0.04 - 0.50 | 0.62 (0.31-0.81) | -1.08 - 0.90 | 88% |
| Chinese centres, with CA125 | 0.30 (0.2-0.41) | 0.08 - 0.60 | 0.50 (0.15-0.71) | -1.10 - 0.86 | 85% |
| European centres, with CA125 | 0.30 (0.22-0.38) | 0.05 - 0.68 | 0.49 (0.37-0.58) | -0.23 - 0.73 | 96% |
| European centres, without CA125 | 0.32 (0.23-0.43) | 0.05 - 0.73 | 0.41 (0.23-0.56) | -0.74 - 0.76 | 86% |
| Non-oncology centres, with CA125 | 0.18 (0.1-0.29) | 0.01 - 0.48 | 0.51 (0.07-0.75) | -1.78 - 0.83 | 86% |
| Non-oncology centres, without CA125 | 0.18 (0.08-0.32) | 0.01 - 0.55 | 0.50 (-0.04-0.78) | -1.98 - 0.85 | 85% |
| Oncology centres, with CA125 | 0.32 (0.26-0.38) | 0.10 - 0.65 | 0.50 (0.39-0.59) | -0.27 - 0.78 | 94% |
| Oncology centres, without CA125 | 0.32 (0.24-0.41) | 0.06 - 0.70 | 0.47 (0.31-0.60) | -0.57 - 0.80 | 88% |
| Postmenopausal, with CA125 | 0.42 (0.32-0.52) | 0.07 - 0.82 | 0.39 (0.20-0.54) | -1.28 - 0.77 | 83% |
| Postmenopausal, without CA125 | 0.44 (0.35-0.54) | 0.11 - 0.78 | 0.34 (0.09-0.53) | -1.21 - 0.73 | 79% |
| Premenopausal, with CA125 | 0.2 (0.14-0.26) | 0.03 - 0.59 | 0.58 (0.45-0.69) | -0.27 - 0.84 | 94% |
| Premenopausal, without CA125 | 0.19 (0.11-0.29) | 0.01 - 0.70 | 0.57 (0.35-0.72) | -1.47 - 0.83 | 87% |

CrI, credible interval; PI, prediction interval

[a] CrI estimated using Bayesian sampling methods

[b] PI calculated using trivariate meta-analysis as in Wynants et al [11]

[c] P(useful) is the probability that the Net Benefit of using the model in a new random centre at the 10% threshold is superior to those of the baseline strategies of treating all or treating none of the patients.

**Table S10. Descriptive data for the meta-analysis of pairwise area under the receiver operating characteristic curve (AUC) calculated with conditional risk method in operated patients.**

| **Meta-analysis** | **Studies** | **Centres** | **Countries** | **N first group** | **N second group** | **TRIPOD adherence** | **References** |
|---|---|---|---|---|---|---|---|
| *ADNEX with CA125* | | | | | | | |
| Benign vs Borderline | 5 | 23 | 9 | 2879 | 295 | 83% | **[17,32,33,41,63]** |
| Benign vs Stage I | 5 | 23 | 9 | 2879 | 306 | 83% | **[17,32,33,41,63]** |
| Benign vs Stage II-IV | 5 | 23 | 9 | 2879 | 644 | 83% | **[17,32,33,41,63]** |
| Benign vs Metastatic | 4 | 22 | 8 | 2583 | 186 | 85% | **[17,32,33,63]** |
| Borderline vs Stage I | 5 | 23 | 9 | 295 | 306 | 83% | **[17,32,33,41,63]** |
| Borderline vs Stage II-IV | 5 | 23 | 9 | 295 | 644 | 83% | **[17,32,33,41,63]** |
| Borderline vs Metastatic | 4 | 22 | 8 | 284 | 186 | 85% | **[17,32,33,63]** |
| Stage I vs Stage II-IV | 5 | 23 | 9 | 306 | 644 | 83% | **[17,32,33,41,63]** |
| Stage I vs Metastatic | 4 | 22 | 8 | 286 | 186 | 85% | **[17,32,33,63]** |
| Stage II-IV vs Metastatic | 4 | 22 | 8 | 619 | 186 | 85% | **[17,32,33,63]** |
| *ADNEX without CA125* | | | | | | | |
| Benign vs Borderline | 4 | 22 | 9 | 2446 | 269 | 88% | **[17,32,41,63]** |
| Benign vs Stage I | 4 | 22 | 9 | 2446 | 267 | 88% | **[17,32,41,63]** |
| Benign vs Stage II-IV | 4 | 22 | 9 | 2446 | 586 | 88% | **[17,32,41,63]** |
| Benign vs Metastatic | 3 | 21 | 8 | 2150 | 165 | 92% | **[17,32,63]** |
| Borderline vs Stage I | 4 | 22 | 9 | 269 | 267 | 88% | **[17,32,41,63]** |
| Borderline vs Stage II-IV | 4 | 22 | 9 | 269 | 586 | 88% | **[17,32,41,63]** |
| Borderline vs Metastatic | 3 | 21 | 8 | 258 | 165 | 92% | **[17,32,63]** |
| Stage I vs Stage II-IV | 4 | 22 | 9 | 267 | 586 | 88% | **[17,32,41,63]** |
| Stage I vs Metastatic | 3 | 21 | 8 | 247 | 165 | 92% | **[17,32,63]** |
| Stage II-IV vs Metastatic | 3 | 21 | 8 | 561 | 165 | 92% | **[17,32,63]** |

Pairwise analysis including metastatic tumours includes 1 study less because **[41]** had too few metastases.
TRIPOD, Transparent reporting of a multivariable prediction model for individual prognosis or diagnosis

**Table S11. Meta-analysis results of pairwise area under receiver operating characteristic curve (AUC) with conditional risk method in operated patients.**

| Meta-Analysis | Studies | Summary estimate | 95% CI | 95% PI | $\tau^2$ |
|---|---|---|---|---|---|
| *ADNEX with CA125* | | | | | |
| Benign vs Borderline | 5 | 0.86 | 0.83 to 0.90 | 0.80 to 0.93 | 0.06 |
| Benign vs Stage I | 5 | 0.92 | 0.88 to 0.96 | 0.82 to >.99 | 0.34 |
| Benign vs Stage II-IV | 5 | 0.98 | 0.97 to 0.99 | 0.96 to >.99 | 0.18 |
| Benign vs Metastatic | 4 | 0.95 | 0.92 to 0.98 | 0.90 to >.99 | 0.22 |
| Borderline vs Stage I | 5 | 0.72 | 0.61 to 0.82 | 0.49 to 0.92 | 0.24 |
| Borderline vs Stage II-IV | 5 | 0.90 | 0.86 to 0.94 | 0.82 to 0.97 | 0.12 |
| Borderline vs Metastatic | 4 | 0.87 | 0.80 to 0.93 | 0.74 to 0.97 | 0.18 |
| Stage I vs Stage II-IV | 5 | 0.82 | 0.75 to 0.88 | 0.68 to 0.94 | 0.16 |
| Stage I vs Metastatic | 4 | 0.78 | 0.70 to 0.85 | 0.64 to 0.90 | 0.10 |
| Stage II-IV vs Metastatic | 4 | 0.78 | 0.71 to 0.85 | 0.65 to 0.90 | 0.09 |
| *ADNEX without CA125* | | | | | |
| Benign vs Borderline | 4 | 0.86 | 0.81 to 0.91 | 0.77 to 0.95 | 0.10 |
| Benign vs Stage I | 4 | 0.92 | 0.88 to 0.97 | 0.82 to >.99 | 0.33 |
| Benign vs Stage II-IV | 4 | 0.97 | 0.95 to 0.98 | 0.94 to 0.99 | 0.17 |
| Benign vs Metastatic | 3 | 0.94 | 0.91 to 0.99 | 0.87 to >.99 | 0.46 |
| Borderline vs Stage I | 4 | 0.73 | 0.60 to 0.85 | 0.48 to 0.94 | 0.29 |
| Borderline vs Stage II-IV | 4 | 0.89 | 0.84 to 0.93 | 0.79 to 0.96 | 0.12 |
| Borderline vs Metastatic | 3 | 0.89 | 0.81 to 0.96 | 0.77 to >.99 | 0.30 |
| Stage I vs Stage II-IV | 4 | 0.75 | 0.63 to 0.85 | 0.51 to 0.95 | 0.30 |
| Stage I vs Metastatic | 3 | 0.78 | 0.68 to 0.90 | 0.61 to 0.96 | 0.22 |
| Stage II-IV vs Metastatic | 3 | 0.66 | 0.50 to 0.81 | 0.39 to 0.90 | 0.27 |

Pairwise analysis including metastatic tumours includes 1 study less because one study **[41]** had too few metastases.
CI, confidence interval; PI, prediction interval

### FIGURES

**Figure S1. TRIPOD (Transparent Reporting of a multivariable prediction model for Individual Prognosis Or Diagnosis) adherence per study (N = 47).**

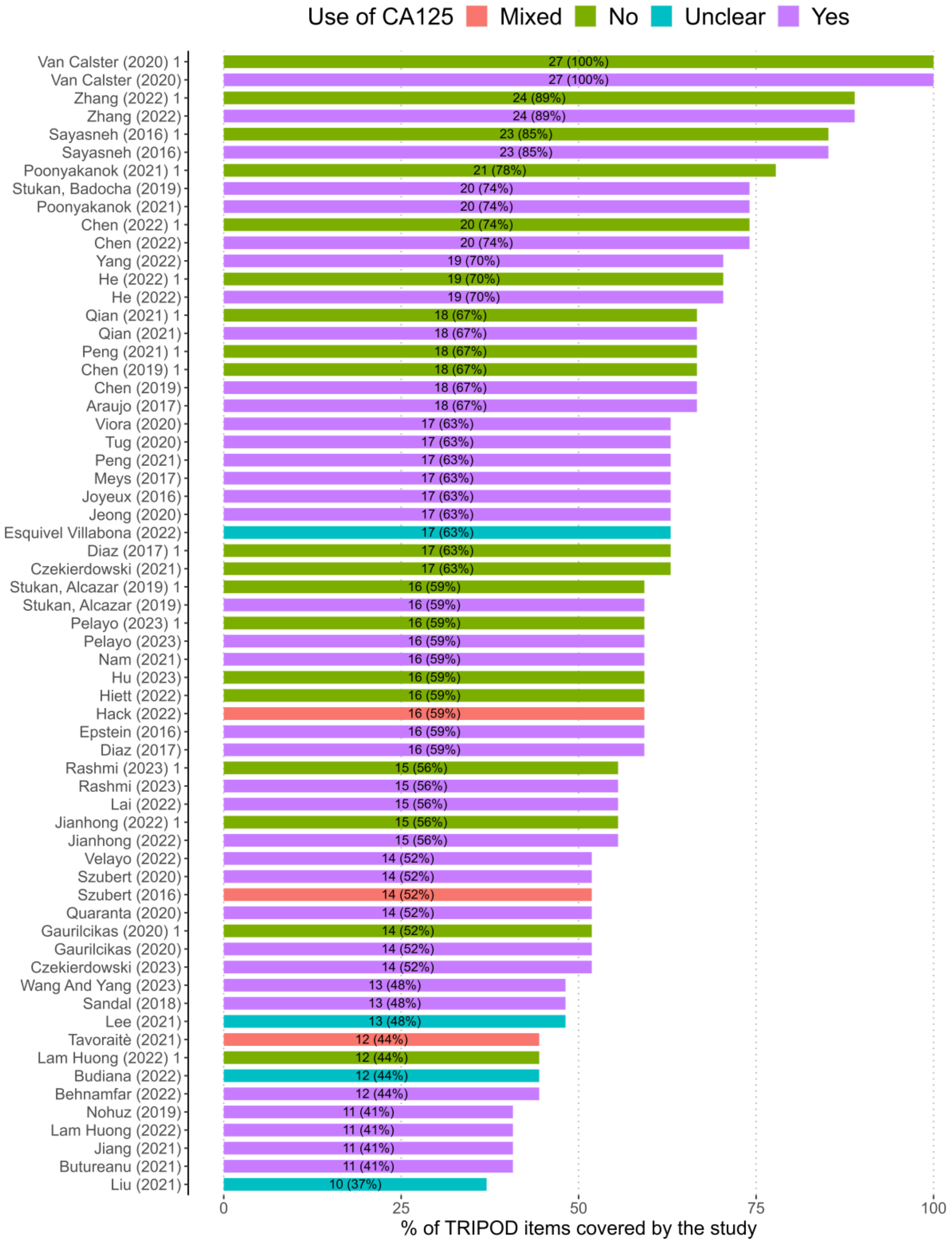

**Figure S2. PROBAST (Prediction model study Risk Of Bias ASsessment Tool) results by subdomain and overall for studies evaluating ADNEX without CA125. Figure generated adapting code from [73]. Blue rows refer to studies included in meta-analysis for the AUC and green rows refer to studies that are included in all meta-analysis.**

| | Participants | Predictors | Outcome | Analysis | Overall |
|---|---|---|---|---|---|
| Chen (2022) | X | + | + | X | X |
| Chen (2019) | - | - | + | X | X |
| Czekierdowski (2021) | X | X | + | X | X |
| Diaz (2017) | X | - | - | X | X |
| Gaurilcikas (2020) | X | X | - | X | X |
| He (2022) | + | + | + | X | X |
| Hiett (2022) | X | + | - | X | X |
| Jianhong (2022) | - | + | - | X | X |
| Lam Huong (2022) | X | + | - | X | X |
| Peng (2021) | X | + | + | X | X |
| Poonyakanok (2021) | - | + | + | X | X |
| Qian (2021) | X | + | + | X | X |
| Zhang (2022) | - | + | + | + | - |
| Hu (2023) | X | + | + | X | X |
| Pelayo (2023) | X | + | + | X | X |
| Rashmi (2023) | + | - | - | X | X |
| Sayasneh (2016) | + | + | + | + | + |
| Stukan, Alcazar (2019) | X | - | - | X | X |
| Van Calster (2020) | + | + | + | + | + |

Judgement
- X High
- \- Unclear
- \+ Low

**Figure S3. PROBAST (Prediction model study Risk Of Bias ASsessment Tool) results by subdomain and overall for studies evaluating ADNEX with CA125. Figure generated adapting code from [73]. Yellow rows refer to studies that are included in meta-analysis for specificity, sensitivity and clinical utility, blue rows refer to studies included in meta-analysis for the AUC and green rows refer to studies that are included in all meta-analysis.**

| | Participants | Predictors | Outcome | Analysis | Overall |
|---|---|---|---|---|---|
| Araujo (2017) | X | - | + | X | X |
| Behnamfar (2022) | X | + | + | X | X |
| Butureanu (2021) | X | - | - | X | X |
| Chen (2022) | X | - | + | X | X |
| Chen (2019) | - | - | + | X | X |
| Czekierdowski (2023) | X | - | X | X | X |
| Diaz (2017) | X | - | - | X | X |
| Gaurilcikas (2020) | X | X | - | X | X |
| Hack (2022) | X | - | - | X | X |
| He (2022) | + | + | + | X | X |
| Jeong (2020) | X | + | X | X | X |
| Jiang (2021) | X | - | - | X | X |
| Jianhong (2022) | - | + | - | X | X |
| Joyeux (2016) | X | - | + | X | X |
| Lai (2022) | X | - | - | X | X |
| Lam Huong (2022) | X | - | - | X | X |
| Nam (2021) | X | - | - | X | X |
| Nohuz (2019) | X | - | - | X | X |
| Peng (2021) | X | - | + | X | X |
| Poonyakanok (2021) | - | - | + | X | X |
| Qian (2021) | X | + | + | X | X |
| Quaranta (2020) | X | - | - | X | X |
| Sandal (2018) | X | - | - | X | X |
| Stukan, Badocha (2019) | - | + | + | X | X |
| Szubert (2020) | X | - | - | X | X |
| Tavoraitè (2021) | X | - | - | X | X |
| Tug (2020) | - | - | - | X | X |
| Velayo (2022) | X | + | - | X | X |
| Viora (2020) | X | - | - | X | X |
| Yang (2022) | - | - | + | X | X |
| Zhang (2022) | - | + | + | + | - |
| Pelayo (2023) | X | + | + | X | X |
| Rashmi (2023) | + | - | - | X | X |
| Wang And Yang (2023) | X | - | - | X | X |
| Szubert (2016) | X | - | - | X | X |
| Epstein (2016) | - | - | - | X | X |
| Sayasneh (2016) | + | + | + | + | + |
| Stukan, Alcazar (2019) | X | - | - | X | X |
| Meys (2017) | X | - | - | X | X |
| Van Calster (2020) | + | + | + | + | + |

Judgement

X High

- Unclear

+ Low

**Figure S4. PROBAST (Prediction model study Risk Of Bias ASsessment Tool) results by subdomain and overall for studies evaluating ADNEX but for which it was unclear whether the version with or without CA125 was used. Figure generated adapting code from [73].**

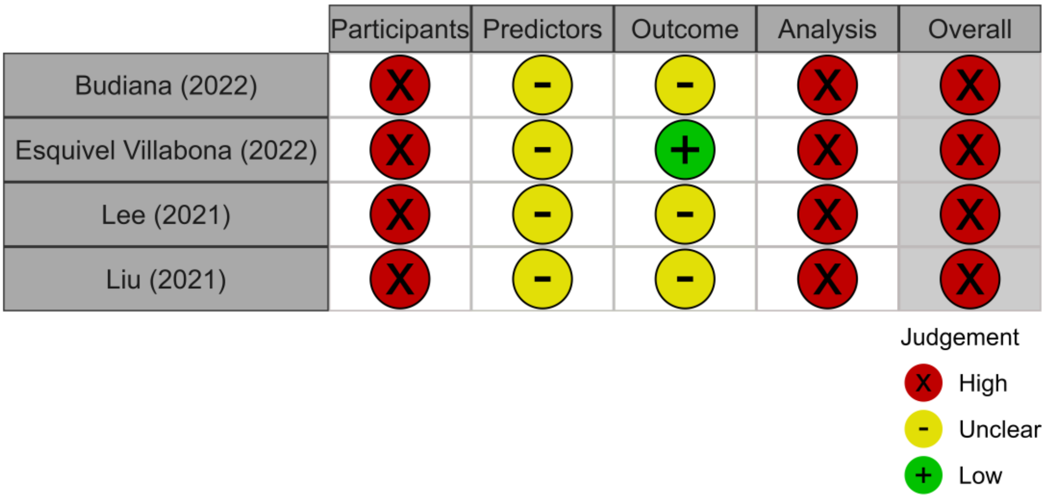

**Figure S5. PROBAST (Distribution of Prediction model study Risk Of Bias ASsessment Tool) results per signalling question in all 63 validations. For PROBAST tool signalling questions see [74,75]. Percentages may sum to 99 or 101 due to rounding.**

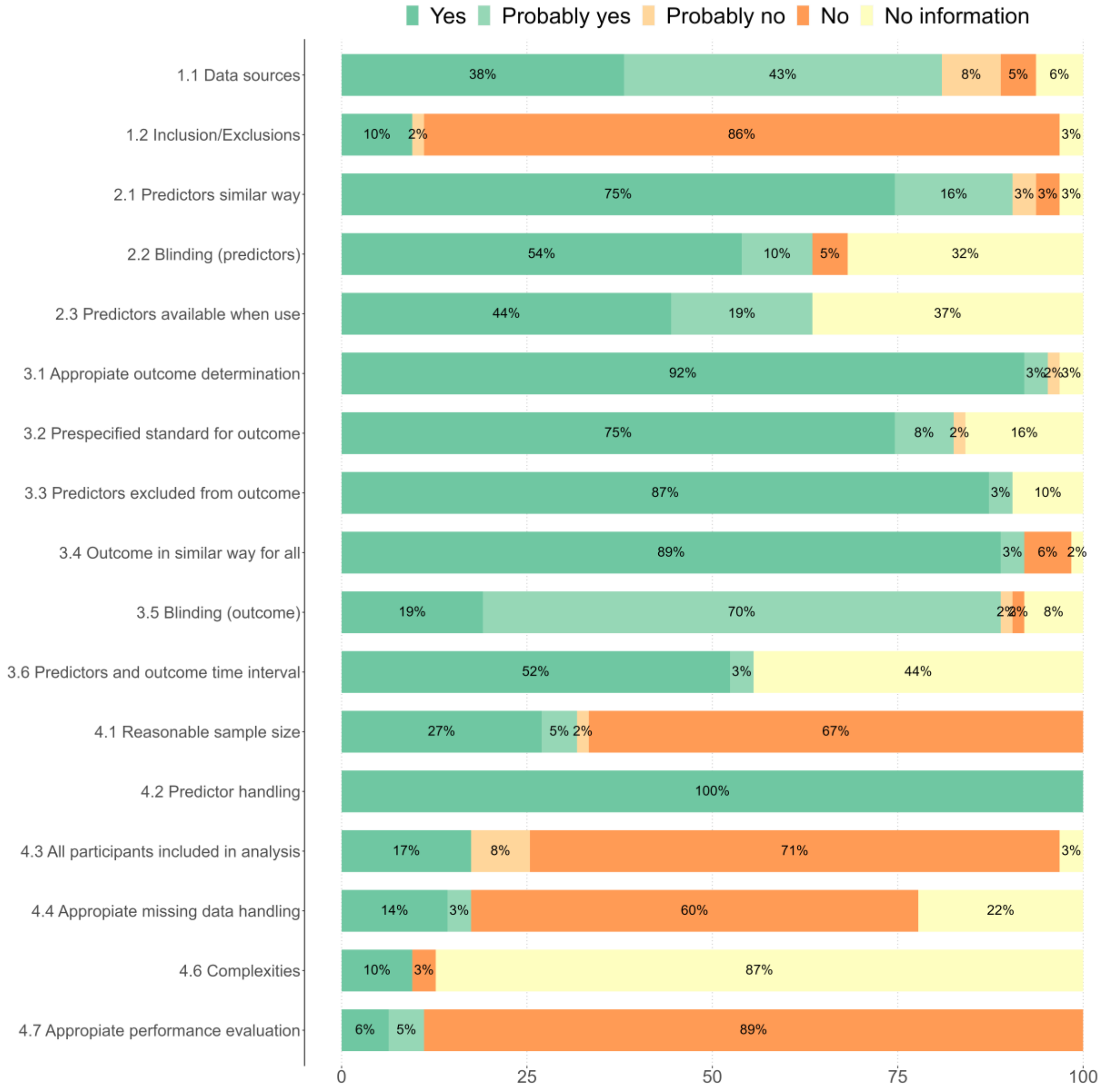

**Figure S6. Forest plot of Net Benefit for ADNEX without CA125. CrI, credible interval; NB, net benefit; Prev, prevalence of malignancy**

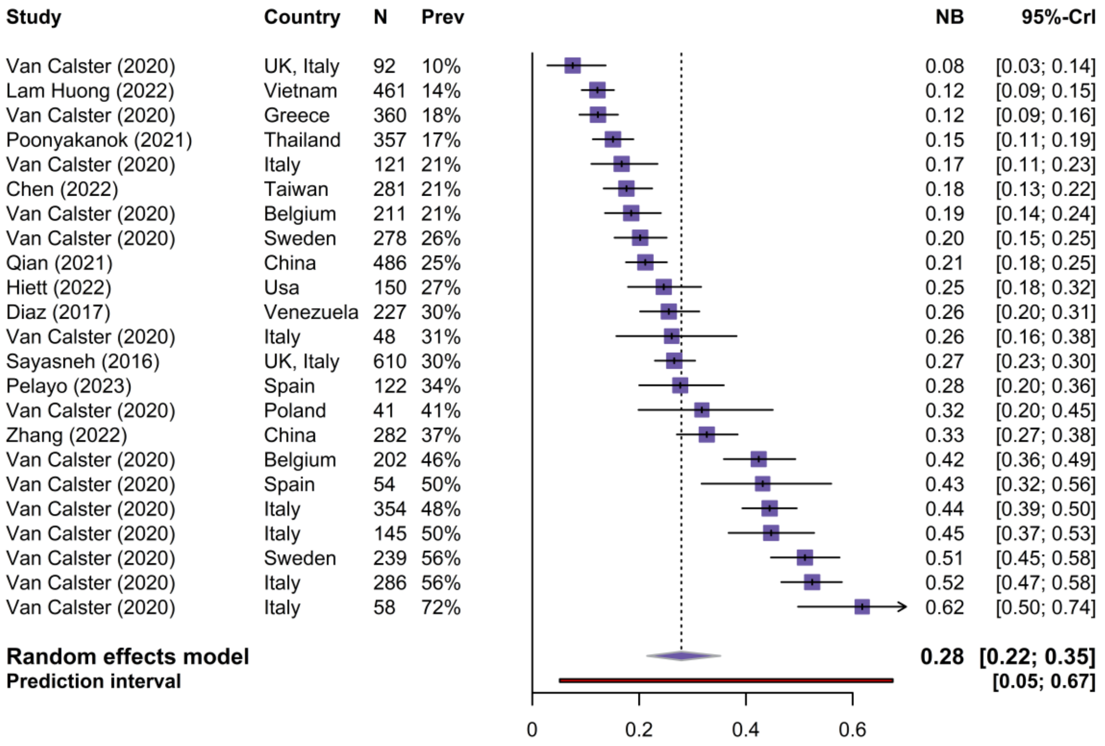

**Figure S7. Forest plot of Relative Utility for ADNEX without CA125. CrI, credible interval; Prev, prevalence of malignancy; RU, relative utility**

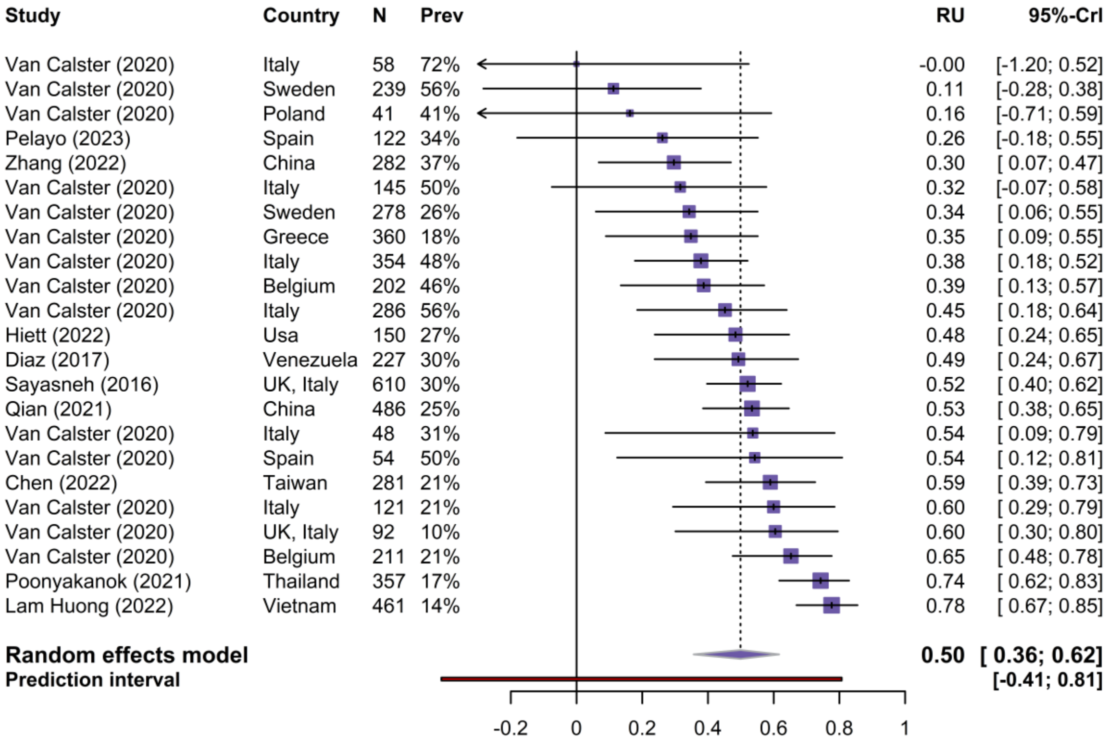

**Figure S8. Forest plot of Net Benefit for ADNEX with CA125. CrI, credible interval; NB, net benefit; Prev, prevalence of malignancy**

| Study | Country | N | Prev | NB | 95%-CrI |
|---|---|---|---|---|---|
| Tug (2020) | Turkey | 285 | 9% | 0.07 | [0.05; 0.11] |
| Van Calster (2020) | UK, Italy | 92 | 10% | 0.08 | [0.04; 0.14] |
| Joyeux (2016) | France | 284 | 11% | 0.09 | [0.05; 0.12] |
| Lam Huong (2022) | Vietnam | 461 | 14% | 0.12 | [0.09; 0.15] |
| Van Calster (2020) | Greece | 360 | 18% | 0.13 | [0.10; 0.17] |
| Poonyakanok (2021) | Thailand | 357 | 17% | 0.16 | [0.12; 0.20] |
| Jeong (2020) | South Korea | 54 | 19% | 0.17 | [0.09; 0.27] |
| Van Calster (2020) | Italy | 121 | 21% | 0.17 | [0.11; 0.24] |
| Chen (2022) | Taiwan | 281 | 21% | 0.18 | [0.13; 0.23] |
| Van Calster (2020) | Belgium | 211 | 21% | 0.18 | [0.13; 0.23] |
| Viora (2020) | Italy | 577 | 25% | 0.21 | [0.18; 0.24] |
| Lai (2022) | China | 734 | 23% | 0.21 | [0.18; 0.24] |
| Qian (2021) | China | 486 | 25% | 0.21 | [0.18; 0.25] |
| Van Calster (2020) | Sweden | 278 | 26% | 0.21 | [0.16; 0.26] |
| Sandal (2018) | Turkey | 191 | 28% | 0.25 | [0.19; 0.31] |
| Szubert (2016) | Spain | 123 | 28% | 0.25 | [0.17; 0.33] |
| Diaz (2017) | Venezuela | 227 | 30% | 0.26 | [0.21; 0.32] |
| Sayasneh (2016) | UK, Italy | 610 | 30% | 0.27 | [0.23; 0.31] |
| Van Calster (2020) | Italy | 48 | 31% | 0.27 | [0.16; 0.39] |
| Yang (2022) | China | 376 | 31% | 0.27 | [0.23; 0.32] |
| Pelayo (2023) | Spain | 122 | 34% | 0.30 | [0.23; 0.38] |
| He (2022) | China | 620 | 35% | 0.30 | [0.27; 0.34] |
| Szubert (2016) | Poland | 204 | 34% | 0.30 | [0.25; 0.37] |
| Meys (2017) | Netherlands | 326 | 35% | 0.32 | [0.27; 0.37] |
| Zhang (2022) | China | 282 | 37% | 0.33 | [0.27; 0.39] |
| Van Calster (2020) | Poland | 41 | 41% | 0.33 | [0.21; 0.46] |
| Wang And Yang (2023) | China | 445 | 40% | 0.37 | [0.32; 0.41] |
| Peng (2021) | China | 224 | 47% | 0.42 | [0.36; 0.49] |
| Van Calster (2020) | Belgium | 202 | 46% | 0.42 | [0.35; 0.49] |
| Van Calster (2020) | Spain | 54 | 50% | 0.44 | [0.32; 0.57] |
| Van Calster (2020) | Italy | 354 | 48% | 0.44 | [0.39; 0.50] |
| Van Calster (2020) | Italy | 145 | 50% | 0.45 | [0.37; 0.52] |
| Stukan, Badocha (2019) | Poland | 100 | 50% | 0.46 | [0.36; 0.56] |
| Araujo (2017) | Brazil | 131 | 52% | 0.46 | [0.38; 0.55] |
| Van Calster (2020) | Sweden | 239 | 56% | 0.51 | [0.45; 0.58] |
| Van Calster (2020) | Italy | 286 | 56% | 0.53 | [0.47; 0.58] |
| Van Calster (2020) | Italy | 58 | 72% | 0.63 | [0.50; 0.74] |
| **Random effects model** | | | | **0.27** | **[0.23; 0.33]** |
| **Prediction interval** | | | | | **[0.06; 0.64]** |

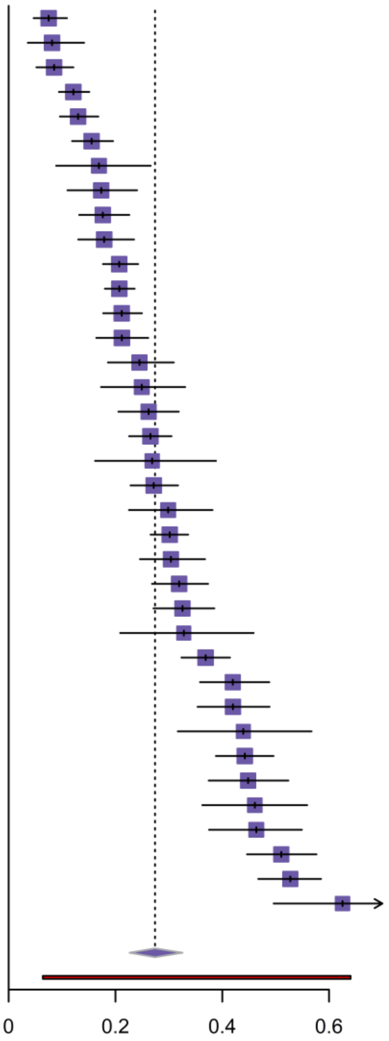

**Figure S9. Forest plot of Relative Utility for ADNEX with CA125. CrI, credible interval; Prev, prevalence of malignancy; RU, relative utility**

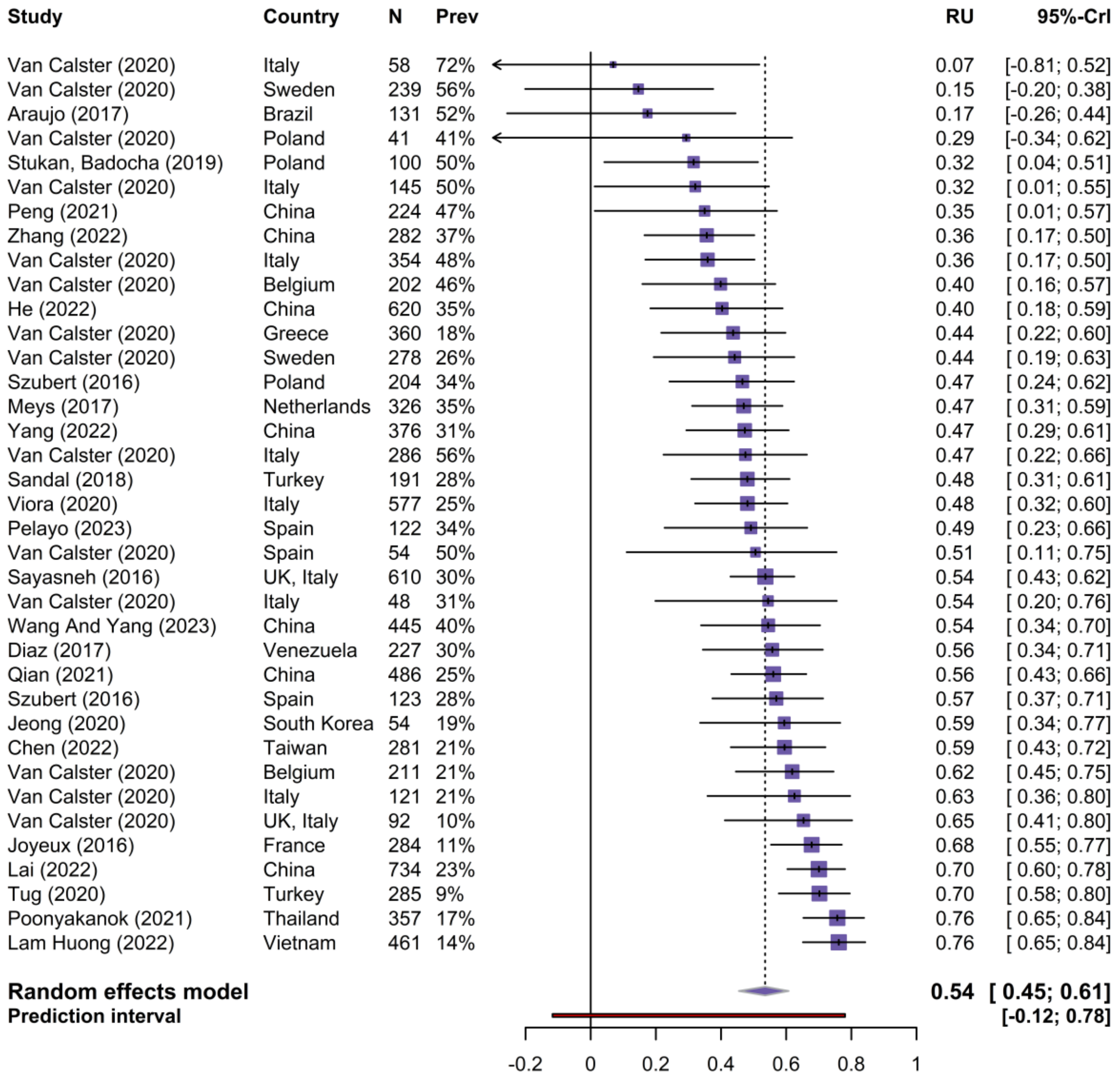

**Figure S10. Meta-regression of the area under the receiver operating characteristic curve (AUC) on the centre-specific prevalence of malignancy for ADNEX with CA 125 (left) and without CA125 (rigt).**

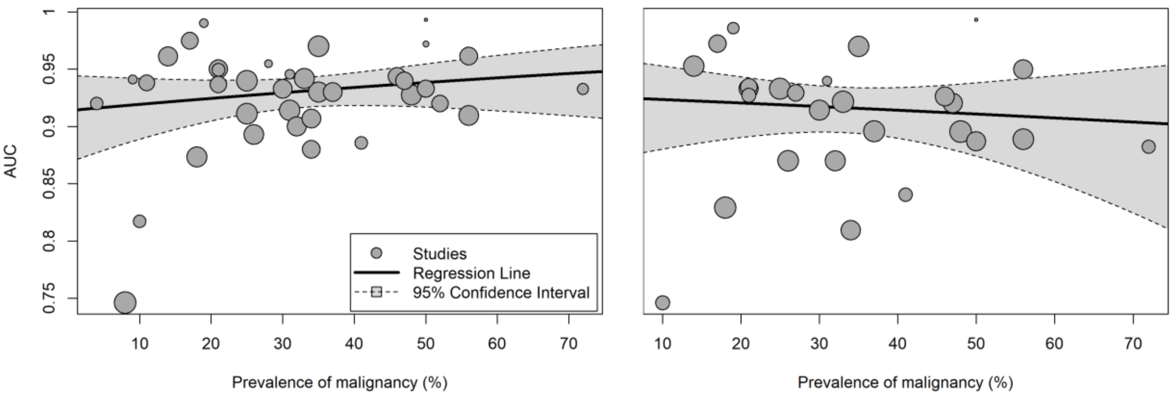

**Figure S11. Meta-regression of sensitivity (left) and specificity (right) on the centre-specific prevalence of malignancy for ADNEX with CA125.**

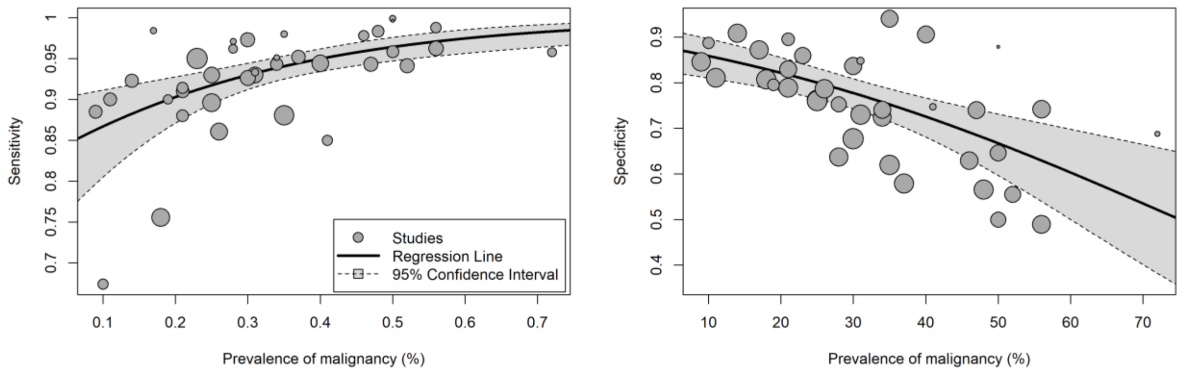

**Figure S12. Meta-regression of sensitivity (left) and specificity (right) on the centre-specific prevalence of malignancy for ADNEX without CA125.**

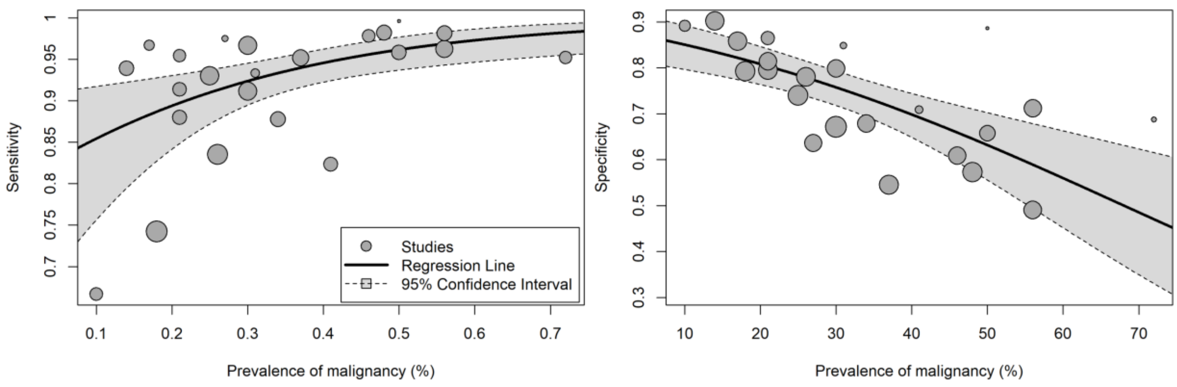

**Figure S13. Funnel plots of the AUC for ADNEX with CA125 validation stratified by prevalence of malignancy (left) and PROBAST Risk of Bias (right).**

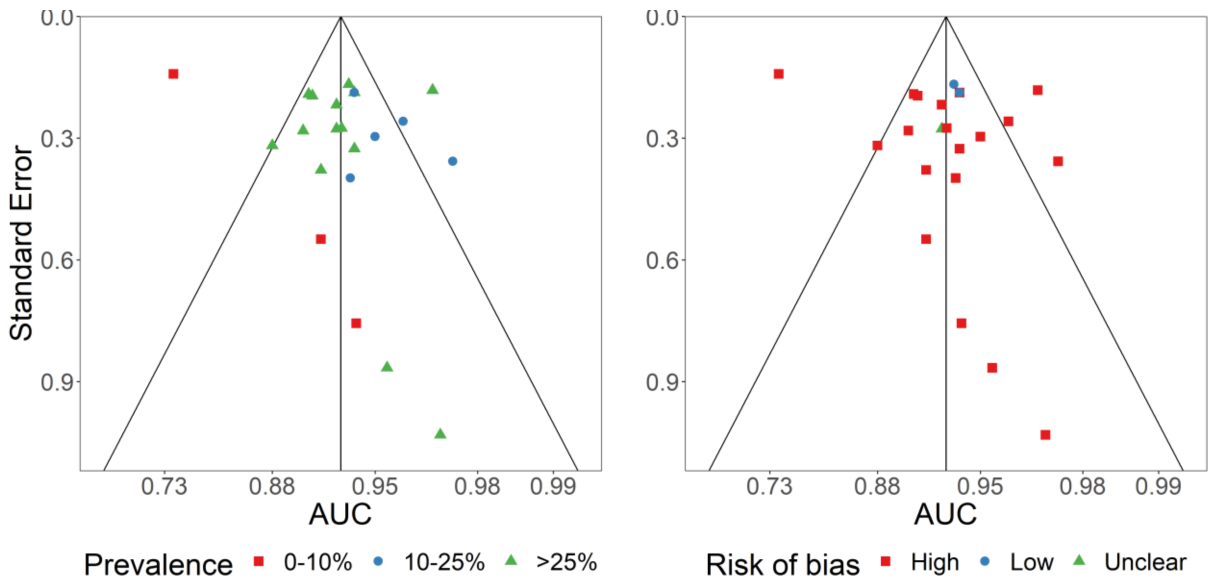
