## Supplementary material for "The ADNEX risk prediction model for ovarian cancer diagnosis: A systematic review and meta-analysis of external validation studies": TRIPOD SRMA Checklist

TRIPOD-SRMA Checklist for reporting systematic reviews of prediction model studies

| **Section and**  **topic** | **Item**  **No** | **Checklist item** | **Page** |
| --- | --- | --- | --- |
| **Title** |  |  |  |
| Title | 1 | Identify the report as a systematic review or meta-analysis (or both) of diagnostic or prognostic model studies. Specify the target population and outcome(s) predicted as relevant to the review question. | Title |
| **Abstract** |  |  |  |
| Abstract | 2 | See the TRIPOD-SRMA Checklist for Abstracts |  |
| **Introduction** |  |  |  |
| Rationale | 3 | Describe the rationale for the review in the context of existing knowledge. | Introduction |
| Objectives | 4 | Provide an explicit statement of the objective(s) being addressed with reference to: target population, index and comparator models (as relevant),  outcome(s), time (prediction horizon and intended moment of using the model), and setting. | Introduction: Last paragraph |
| **Methods** |  |  |  |
| Study eligibility  criteria | 5 | Specify study characteristics used as eligibility criteria, including any prediction models of specific interest, and whether development or validation  studies (or both) were eligible. | Eligibility criteria |
| Information  sources | 6 | Specify all databases, registers, websites, organisations, reference lists and other sources searched or consulted to identify studies. Specify the date  when each source was last searched or consulted. | Information sources and search strategy |
| Search strategy | 7 | Present the full search strategies for all databases, registers and websites, including any filters and limits used. | Supp. Material S1 |
| Study selection  process | 8 | Specify the methods used to decide whether a study met the inclusion criteria of the review, including how many reviewers screened each record  and each report retrieved, whether they worked independently, and if applicable, details of automation tools used in the process. | Study selection |
| Data collection process | 9 | Specify the methods used to collect data from study reports, including how many reviewers collected data from each report, whether they worked independently, any processes for obtaining or confirming data from study investigators, and if applicable, details of automation tools used in the  process. | Data extraction and data items |
| Data Items | 10a | List and define all items for which data were sought from each study. | Data extraction and data items  Supp Table S1  OSF Extraction sheet |
|  | 10b | State the model performance measures that were sought (e.g., measures of calibration, discrimination, overall model fit, clinical utility). | Data extraction and data items  Supp Table S1  OSF Extraction sheet |
|  | 10c | Describe how any desired but unreported data items (items 10a, 10b) were handled (e.g., contacted authors, calculated from other reported  information). | Data extraction and data items |
| Risk of bias and applicability  assessment | 11 | Specify the methods used to assess risk of bias in the included studies and their applicability to the review question. This should be done separately for each model development and validation. Include details of any tool(s) used, how many reviewers assessed each study and whether they worked  independently. | Data extraction and data items |
| Synthesis methods | 12a | Describe any methods for synthesising estimates of performance measures for each model. If meta-analysis was carried out, describe the methods used, including any transformations of data prior to pooling, how any heterogeneity in model performance was quantified and handled, and  software package(s) used. | Statistical analysis and quantitative data synthesis |
|  | 12b | Describe any methods used to explore possible causes of heterogeneity in model performance (e.g., subgroup analysis, meta-regression), including  whether or not they were planned. | Statistical analysis and quantitative data synthesis |

| **Section and**  **topic** | **Item**  **No** | **Checklist item** | **Page** |
| --- | --- | --- | --- |
|  | 12c | Describe any sensitivity analyses conducted to assess robustness of the synthesised results. | Statistical analysis and quantitative data synthesis |
| Certainty  assessment | 13 | Describe any methods used to assess certainty (or confidence) in the body of evidence for a prediction model. | Statistical analysis and quantitative data synthesis |
| **Results** |  |  |  |
| Study selection | 14 | Describe the results of the search and selection process, from the number of records identified in the search to the number of studies and models included in the review, ideally using a flow diagram. | Results and figure 1 |
| Study and model  characteristics | 15 | Present study characteristics and model details extracted (as per Item 10a), and cite the study reports. | Results  Table S3 |
| Risk of bias and  applicability | 16 | Present results of risk of bias and applicability assessment. This should be done separately for each model development and validation in each  included study. | Figures S2-5 |
| Results of model performance in individual  studies | 17 | Present performance estimates and confidence intervals for each model and all evaluations, including whether they relate to the internal or external validation performance. If internal, give details of the method. | Table S4 and forest plots (Figures 4-7, S6-S9) |
| Results of syntheses | 18a | Present the results of any synthesis of model performance, together with details of which study estimates contributed. If meta-analysis was carried  out, then for each model and performance measure, present summary results, confidence/credible intervals and measures of heterogeneity. Forest plots may be useful. | Tables 2,S6-11 |
|  | 18b | For each model, present results of all investigations of possible causes of heterogeneity in model performance. | Tables 2, S8,S9,S11 |
|  | 18c | Present results of all sensitivity analyses conducted to assess the robustness of the synthesised results. | Tables 2, S8,S9,S11 |
| Certainty of  evidence | 19 | Present any assessments of certainty (or confidence) in the body of evidence for each prediction model of interest. | Results Last paragraph. |
| **Discussion** |  |  |  |
| Summary of  evidence | 20 | Summarise the main findings including the strengths and limitations of the evidence. | Discussion |
| Limitations | 21 | Discuss the strengths and limitations of the review process. | Discussion |
| Implications | 22 | Discuss implications of the results in the context of other evidence and for practice, policy, and future research. | Discussion |
| **Other**  **information** |  |  |  |
| Registration and protocol | 23a | Provide registration information for the review, including register name and registration number, or state that the review was not registered. | Protocol registration |
|  | 23b | Indicate where the review protocol can be accessed, or state that a protocol was not prepared. | Protocol registration |
|  | 23c | Describe and explain any amendments to information provided at registration or in the protocol. | Protocol registration |
| Support | 24 | Describe sources of financial or non-financial support for the review, and the role of the funders or sponsors in the review. | Funding |
| Competing  interests | 25 | Declare any competing interests of review authors. | Conflicts of interest |

| **Section and**  **topic** | **Item**  **No** | **Checklist item** | **Page** |
| --- | --- | --- | --- |
| Availability of  data, code, and other materials | 26 | Report which of the following are publicly available and where they can be found: template data collection forms; data extracted from included studies; data used for all analyses; analytic code; any other materials used in the review. | Availability of data and code |

This checklist appears in appendix 2 of Snell KIE, Levis B, Damen JAA, et al. Transparent reporting of multivariable prediction models for individual prognosis or diagnosis: checklist for systematic reviews and meta-analyses (TRIPOD-SRMA). *BMJ* 2023;381:e073538. doi:10.1136/bmj-2022-073538.
